# Neuroinflammation distinguishes HLA haplotypes in progressive supranuclear palsy

**DOI:** 10.1101/2025.07.21.25331869

**Authors:** Shelley L. Forrest, Sarah S. Zaheer, Ain Kim, Hidetomo Tanaka, Helen Chasiotis, Jun Li, Susan H. Fox, Jinguo Wang, M. Carmela Tartaglia, Anthony E. Lang, Gabor G. Kovacs

## Abstract

**Objectives:** Progressive supranuclear palsy (PSP) is a neurodegenerative 4R tauopathy clinically presenting with atypical parkinsonism or cognitive behavioral changes and a relatively uniform neuropathology. We recently identified rare HLA haplotypes in PSP and now examine whether HLA haplotypes are associated with different cytopathological and clinical phenotypes.

**Methods:** Retrospective collection of clinical data and mapping of T and B cells, microglia, and phosphorylated-tau (p-Tau) cytopathologies in 32 PSP cases. Machine learning was used to analyze whether pathological variables and their ratios, or the sequence of clinical symptoms cluster or predict HLA haplotypes.

**Results:** Four groups were defined based on HLA haplotypes: i) 12 cases with the haplotype associated with narcolepsy (*DRB1*\*15:01-*DQB1*\*06:02); ii) 11 cases with other *DQ5-DQ6* haplotypes; iii) 8 cases with various haplotypes frequent in the general population; and iv) one case with the haplotype frequent in IgLON5-disease (*DRB1*\*10:01-*DQB1*\*05:01). Neuropathology revealed regional differences in the severity of microglia load, density of cytotoxic T cells, and p-Tau cytopathologies between groups. HLA haplotypes were most distinguishable using machine learned features of inflammatory markers and ratios of neuropathological variables (clustering accuracy: 86.96% and 91.30%, respectively). The sequence of clinical symptoms and the ratios of neuropathological variables were the strongest predictors of HLA haplotypes (prediction accuracy=80.00% and 71.43%, respectively).

**Interpretation:** PSP pathology might be associated with various etiological-pathogenic events including targetable autoimmune mechanisms. The HLA-haplotype dependent diversity of neuroinflammatory markers should be evaluated in clinical and biomarker studies in, and beyond, PSP to understand its relevance for patient stratification in disease modifying therapy trials.

## Introduction

Progressive supranuclear palsy (PSP) is a neurodegenerative 4R tauopathy associated with the accumulation of phosphorylated tau (p-tau) in neurons, oligodendrocytes, and astrocytes.^1^ PSP is considered a homogenous disorder based on hallmark neuropathological features^1^ and similar filament structure of misfolded tau reported in six PSP cases examined to date.^2^ However, clinical phenotypes associated with PSP pathology vary and include syndromes in the spectrum of atypical parkinsonism and cognitive behavioral changes.^3^ Clinical variability is associated with differences in the distribution patterns of p-tau cytopathologies^4^ and in cases with Richardson syndrome, a six-tiered neuropathological staging system was introduced.^4^

Although neurodegenerative diseases are traditionally defined by neuronal loss not attributable to peripheral neoplastic or inflammatory cell infiltration (i.e. as in infectious encephalitis) or microbiological or environmental toxic agents,^5^ the concept of neuroinflammation in neurodegenerative diseases is frequently emphasized with respect to biomarker and therapy development.^6^ While the term neuroinflammation has been used in various, and not necessarily harmonized settings, currently the most accepted definition is that it constitutes the activation of microglia and astrocytes and the accumulation of infiltrated leukocytes as the body’s attempt to reduce injury and promote repair.^6^ Importantly, PSP-type pathology overlaps with some neuropathological features described in IgLON5 autoimmune encephalitis-related tauopathy^7^ and the 3R and 4R tauopathy, postencephalitic parkinsonism.^8^ In the original paper conceptualizing PSP as a novel disease entity, care was taken to distinguish it from postencephalitic parkinsonism.^9^ Indeed, while plasma cells were not observed, some level of lymphocytic cuffing of blood vessels was described.^9^ Interestingly, recent studies using immunostainings for lymphocytes have shown that CD8 cytotoxic T cells are consistently present in the midbrain in PSP, and are more prominent than observed in other neurogenerative parkinsonisms.^10, 11^ These autopsy studies were complemented by bodily fluid studies such as cytokine profiles^12, 13^ or shift in peripheral CD4 and CD8 positive T cell populations.^13^

However, in all the aforementioned studies, variability between PSP cases was observed for the neuroinflammation markers examined. Based on the fact that the human leukocyte antigen (*HLA*) locus on chromosome 6 has been implicated in several autoimmune disorders and plays an important role in the adaptive immune system,^14^ we hypothesized that one reason could be that PSP cases have different HLA haplotypes. Indeed, our recent study identified the *DRB1*\*15:01-*DQB1*\*06:02 haplotype and *DQB1*06:01* and *DQB1*\*06:02 alleles are strongly associated with PSP and identified potential epitopes within the tau peptide that may bind to alleles overexpressed in PSP patients. Importantly, this raises the possibility that a subset of PSP cases might have an autoimmune pathophysiological component.^15^ Therefore, in the present study we performed systematic mapping of p-tau pathology and inflammatory markers in the brains of PSP cases stratified based on the HLA haplotypes.

## Material and Methods

### Clinical information

Clinical data was analyzed from a cohort of 32 patients clinically diagnosed with PSP, comprising 11 retrospective cases and 21 prospective cases obtained from the Rossy PSP Centre at the University Health Network (UHN). Demographic variables included sex and age at death, which encompassed both natural death and death via Medical Assistance in Dying (MAiD). Disease duration was calculated from the time of onset of the first reported symptom to the time of death. For this study, we focused on symptoms explicitly documented in the clinical records. Symptom chronology was assessed by identifying the first, second, and third reported symptoms in the order of their appearance. The presence of each symptom was annotated as “+” if reported, “-” if the symptom was not reported, tested and found absent, and “N/A” if the data were unavailable. To further characterize disease severity and clinical heterogeneity, we recorded additional features commonly associated with PSP, including vertical supranuclear gaze palsy, bulbar dysfunction, gait dysfunction and presence of falls, parkinsonian features, psychiatric manifestations, cognitive dysfunction and behavioral disturbances, language abnormalities, and sleep disturbances.

### Neuropathology

Autopsy tissue from human brains was collected with informed consent from patients or their relatives and approval by the University Health Network Research Ethics Board (Nr. 20-5258). A systematic neuropathological examination was performed following the established diagnostic criteria.^16, 17^ For all cases HLA genotyping was available as recently reported.^15^ For the evaluation of inflammatory markers we included brain regions affected in early and moderate stages of PSP, including midbrain (oculomotor complex, substantia nigra, cerebral peduncle, red nucleus and superior cerebellar peduncle), posterior portion of basal ganglia (globus pallidus, putamen, internal capsule), thalamus area (subthalamic nucleus and medial thalamic nucleus), and motor cortex (gray and white matter separately). A four-tiered (none, mild, moderate, severe) semiquantitative scoring approach was used to describe p-tau cytopathologies (neuronal, astrocytic, and oligodendrocytic). Heatmaps were generated by standardizing the data using z-score and converting the quantification of marker-based immunoreactivity into quartile values, where 0 = no immunoreactivity, 1 = low, 2 = intermediate, 3 = high, and 4 = very high, and these scores were then indicated on the heatmap template using a color scale bar (score 0 = white, score 1 = light yellow, score 2 = yellow, score 3 = orange, score 4 = red). Regions not evaluated are indicated in grey.

### Immunohistochemistry

Immunohistochemistry was performed using the following primary antibodies: anti-phospho-tau (clone AT8, pSer202/Thr205, 1:1000, Thermo Fisher Scientific), microglia marker anti-HLA DR + DP + DQ (clone CR3/43, 1:200, Abcam), anti-CD3 (1:200, polyclonal, Dako/Agilent), anti-CD4 (clone EPR6855, 1:500, Abcam), anti-CD8 (clone C8/144B, 1:200, Dako/Agilent), and anti-CD20 (clone L26, 1:500, Dako/Agilent). In each case 6 immunostainings were performed in 4 paraffin embedded tissue blocks (in sum 768 sections). Dako PT Link with low pH solution was used for antigen retrieval for all antibodies and then immunostaining was performed using the Dako Autostainer Link 48 and EnVision FLEX+ Visualization System, according to the manufacturer’s instructions. All sections were subsequently counterstained with haematoxylin. Stained sections were digitized using the Huron TissueScope LE120 slide scanner, and images were imported into HALO software (Indica Labs) for manual annotation of subregions of interest (11 anatomical subregions) and automated quantification of AT8, microglia, CD3, CD4, CD8 and CD20 area cell density within the annotated subregions of interest. Since a few subregions were not identifiable, in summary this resulted in 1954 annotated subregions. Neuromelanin was manually excluded from analysis in the substantia nigra. Detailed procedures used in HALO analysis are provided in the **online Supplementary file Methods and Tables S1, S2, and Figures S1-3)**. For the semiquantitative evaluation of p-Tau cytopathologies we used a six-tiered approach as follows: none (0), single (0.5), mild (1), moderate (2), severe (3), extremely severe (3.5).

### Statistical analysis

Fisher’s exact test was used to compare the frequency of presenting clinical symptoms. Kruskal-Wallis and Mann-Whitney-U test was performed to compare total p-tau load, microglia load and CD8, CD4, CD3, CD20 cell counts, ratios of neuropathological variables, duration of illness, and age at death between groups and between regions in the same groups. Student’s T-test was used to compare semiquantitative p-tau cytopathology scores between groups and brain regions. Spearman correlation test was used to compare association between pathological variables, duration of illness, and age at death. To correct for the effect of PSP neuropathological stage,^4^ Alzheimer’s disease neuropathologic change (ADNC) level,^18^ Lewy body disease (LDB) stage,^19^ duration of illness, and age at death on the differences between groups we used the non-parametric analysis of covariance (ANCOVA).

### Machine learning analysis

Four separate datasets were analyzed: quantification of inflammatory marker-based pathology (i.e., microglia, p-Tau, CD3, CD4, CD8 and CD20), quantification of cytopathology (i.e., neuronal-, astrocytic- and oligodendrocytic-tau), sequence of clinical symptoms (i.e., first, second and third symptoms) and the ratios of the inflammatory marker-based pathology (i.e., CD20/CD8, p-Tau/CD8, AT8/CD4, microglia/CD8, microglia/CD4 and CD8/CD4).

For all datasets, an unsupervised Gaussian Mixture Model (GMM) was used to evaluate whether the data would naturally cluster into the three haplotypes, given all the features in the dataset. Subsequently, one-hot encoded leaf indices were extracted from a supervised Random Forest Classifier (RFC) and were used to perform unsupervised GMM on the extracted features. The RFC-derived, one-hot encoded leaf indices transform the raw features into high-dimensional embedding space, allowing for improved unsupervised clustering that captures class-relevant structures. The adjusted rand score (ARI) and clustering accuracy after applying the Hungarian algorithm to align the GMM cluster labels with the true HLA haplotype labels were reported to assess clustering performance. To evaluate predictability, a supervised Random Forest Classifier (RFC) was used to predict the true HLA haplotypes, based on the features in each dataset and feature extraction was used to identify important features that distinguish the different HLA haplotypes. For the progression of clinical symptoms, where 1^st^, 2^nd^ and 3^rd^ symptoms were indicated with a category (0 = none, 1 = gait instability including falls, 2 = cognitive/behavioral, 3 = oculomotor, 4 = slowness or stiffness, 5 = early combination of gait instability and slowness and 6 = other), one-hot encoding was applied to the dataset to interpret categorical data into numerical data, and Long Short-Term Memory (LSTM) autoencoding was used to learn latent features, prior to any GMM or RFC analysis.

ADNC, PSP stage, duration of illness, and age at death were incorporated in the dataset in the effort to adjust for any confounding effects. Prior to any analysis, all data were standardized, and any missing values were removed. All algorithms used in this study were sourced from the scikit-learn library (https://scikit-learn.org) and visualizations were created using the matplotlib library (https://matplotlib.org) and seaborn (seaborn.pydata.org), using Python 3.11 on Google Colab (https://colab.research.google.com).

## Results

### Grouping of cases

Four groups were defined based on HLA haplotypes:^15^ 1) 12 cases with a haplotype associated with narcolepsy (*DRB1*\*15:01-*DQB1*\*06:02); 2) 11 cases with other *DQ5-DQ6* haplotypes; 3) 8 cases with various haplotypes frequent in the general population; and 4) one case with the haplotype frequent in IgLON5-associated encephalitis (*DRB1*\*10:01-*DQB1*\*05:01).

### Clinical and neuropathological characteristics of HLA-haplotype groups

Age at death (76.11+2.36; 77.22+2.14, and 75.86+2.04; p=0.68) and duration of illness (6.33+0.60, 10.10+2.25, and 8.00+1.34, p= 0.23) of cases with PSP stage > 3 were not significantly different between HLA-haplotype groups 1-3, respectively; furthermore, chi^2^ test did not show differences between the distribution of sexes (*p*=0.69).

Most cases showed PSP stage ≥ 3 p-tau pathology in all groups (**Table 1**); chi^2^ did not show differences between the distribution of stages (*p*=0.60). Group 3 of our cohort consisted of various haplotypes and pathological variants including a case suspected to have postencephalitic parkinsonism showing 3R and 4R tau pathology in the substantia nigra^20^ and group 2 included a case compatible with limbic predominant 4R tauopathy (LNT) type pathology overlapping with PSP type pathology.^2, 21^ None of the co-pathologies (**Table 1**) differed significantly between groups (*p*>0.2 for all). Heatmapping of the mean values of total p-tau load and various inflammatory markers revealed distinct patterns for the different groups as also observed in tissue sections (**Figure 1**).

**Figure 1.**
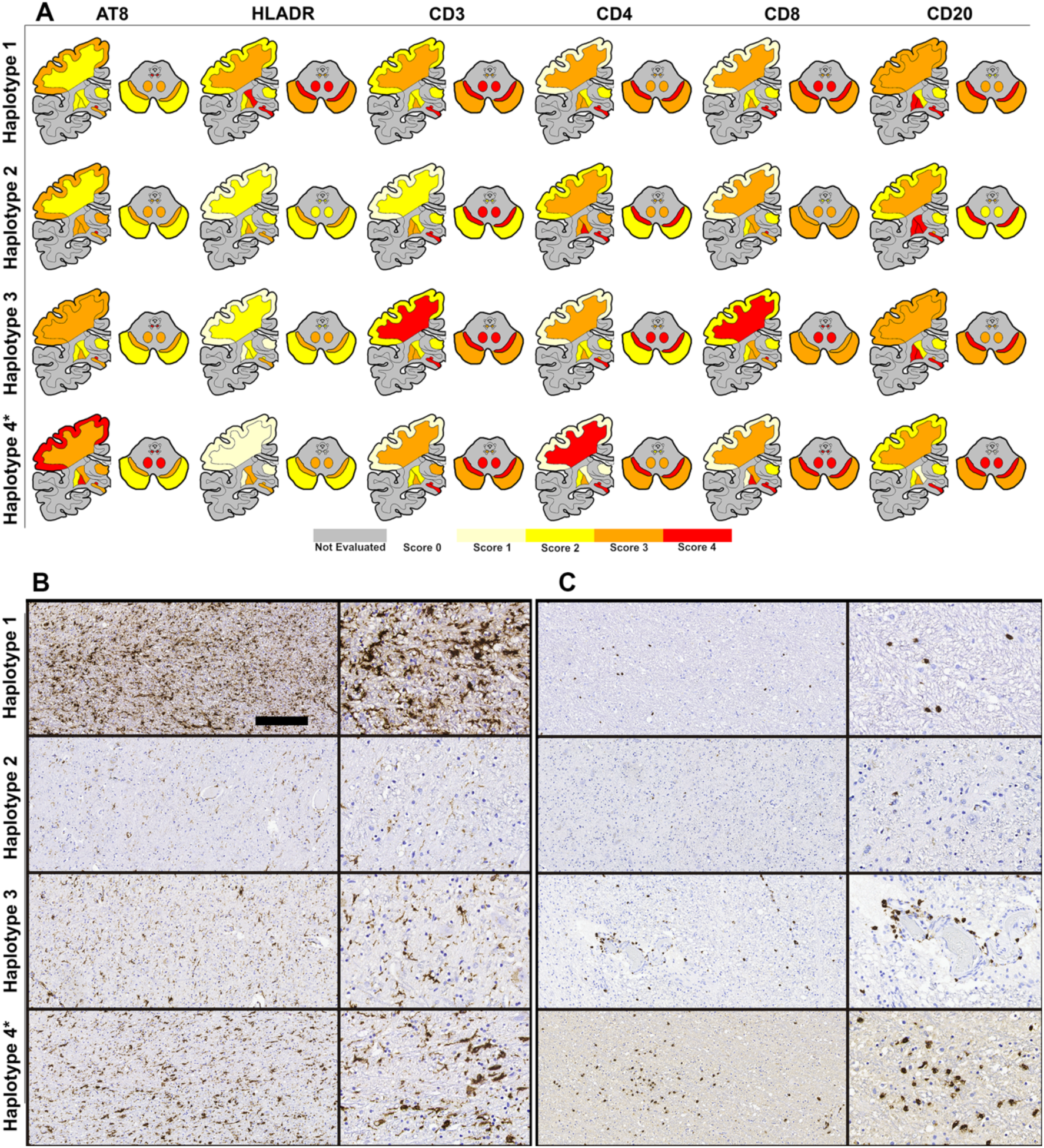
Heatmap of marker-based pathological load in HLA haplotypes. **(A)** Quantification of marker-based immunoreactivity in the globus pallidus, internal capsule, putamen, motor cortex and underlying WM, medial nucleus, subthalamic nucleus, peduncle, substantia nigra, superior cerebellar peduncle and oculomotor complex were converted to scores, where 0 = no immunoreactivity (white), 1 = low (light yellow), 2 = intermediate (yellow), 3 = high (orange), and 4 = very high (red). Regions not evaluated are indicated in grey. Each brain represents the averaged marker-based pathological load in different HLA haplotypes. *Haplotype 4 represents values from a single case only. (**B**) Representative images of the microglia staining (HLA-DR) and (**C**) CD8 immunostaining in the red nucleus/superior cerebellar peduncle area with lower (left side of **B** and **C**) and higher magnification (right side of **B** and **C**). Bar in upper image of **B** indicates 150 μm for the low and 80 μm for the high magnification images.

**Table 1.**
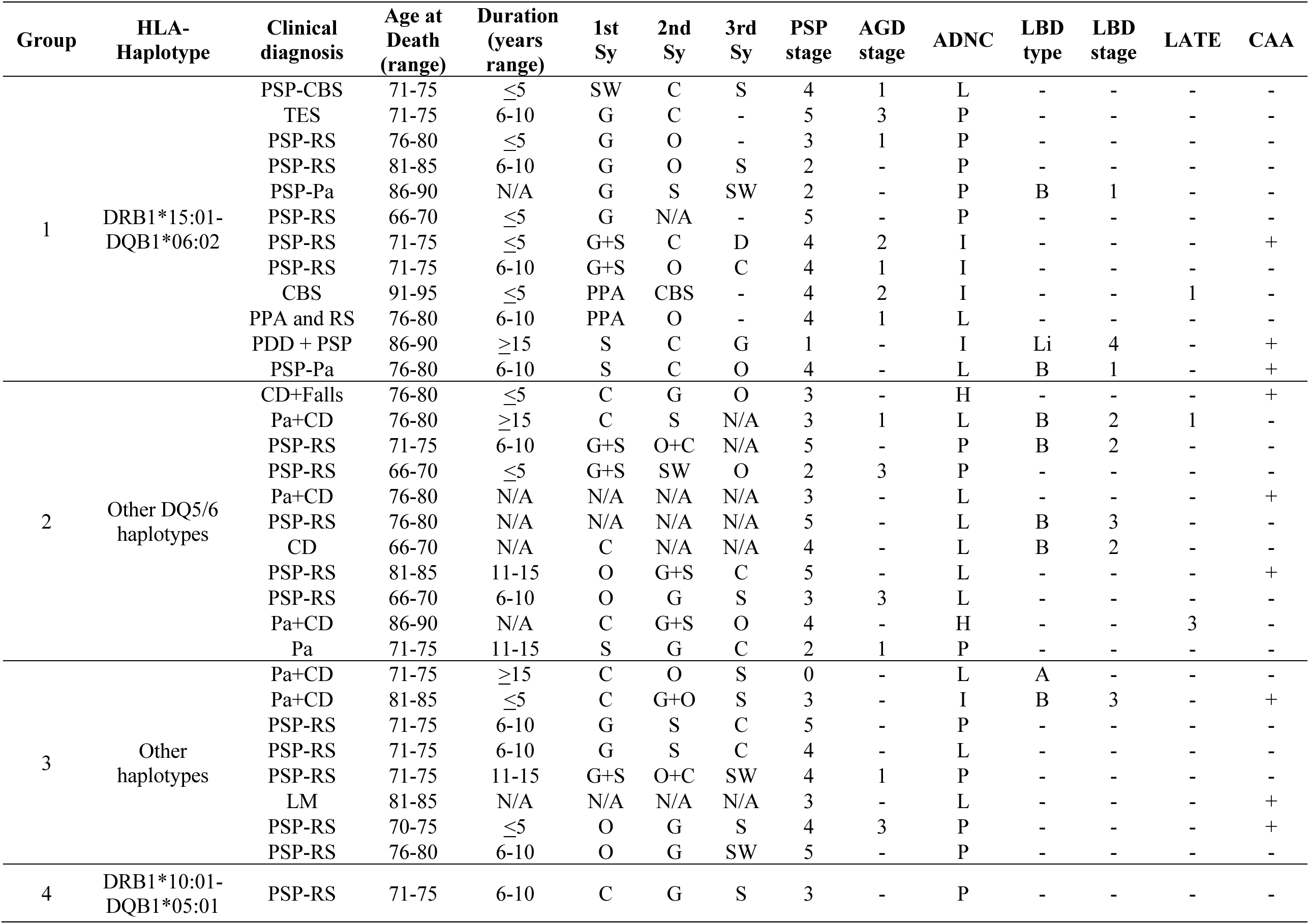
Clinical and neuropathological data of the cohort examined grouped based on HLA haplotypes. **Abbreviations:** A: amygdala, ADNC: Alzheimer’s disease neuropathologic change, AGD: argyrophilic grain disease, B: brainstem, CAA: cerebral amyloid angiopathy, CBS: corticobasal syndrome, C: cognitive, CD: cognitive decline, D: dystonia, F: female, G: gait, H: high, I: intermediate, L: low, Li: limbic, LATE: limbic age-related TDP-43 encephalopathy, LBD: Lewy body disease, LM: limited mobility, M: male, N/A: not available, O: oculomotor, PSP: progressive supranuclear palsy, P: primary age-related tauopathy, Pa: parkinsonism, PDD: Parkinson’s disease dementia, PPA : primary progressive aphasia, RS: Richardson syndrome, S: slowness/stiffness, SW: swallowing difficulty/Slurred speech, TES: traumatic encephalopathy syndrome. +: present, -: not present. PSP stage according to Kovacs et al.,^4^ AGD stage according to Saito et al.,^34^ ADNC according to Montine et al.,^18^ LBD type according to Attems et al.,^35^ LBD stage according to Braak et al.,^19^ LATE according to Nelson et al.^36^

Group 1 cases were more likely to present with gait instability and stiffness compared to group 2, which showed other presenting symptoms, including cognitive/behavioral or oculomotor dysfunction more frequently (*p*=0.08). Group 3 cases showed cognitive, oculomotor dysfunction or gait instability as the presenting symptom. For machine learning algorithms, one case with the haplotype frequently associated with IgLON5-associated encephalitis (*DRB1*\*10:01- *DQB1*\*05:01) and the case with suspected postencephalitic parkinsonism in group 3 was removed from all datasets prior to analysis. Based on the sequence of clinical symptoms, the unsupervised GMM had no meaningful clustering and low clustering accuracy (ARI = -0.0618 and clustering accuracy = 41.67%). However, subsequent unsupervised GMM on the extracted features improved the clustering accuracy (ARI = 0.0881 and clustering accuracy = 54.17%). The prediction accuracy of HLA haplotypes based on the sequence of clinical symptoms was 80.00%. Since the features are LSTM-derived, the top features contributing to the high predictability cannot be identified.

### Increased microglia load is a distinguishing feature of DRB1*15:01-DQB1*06:02 HLA haplotype

Microglia load was higher in several regions in group 1 compared to those in group 2 and 3 (**online Supplementary file Results**). In all regions examined, we did not find significant correlations between PSP stage and microglia load in group 1 (range of *p* values 0.06-0.90) and group 2 (range of *p* values 0.49-0.92). For group 3 cases there was a correlation in the oculomotor complex (rho=0.74, *p*=0.035) and substantia nigra only (rho=0.86, *p*=0.006). Pooling all regions and cases examined (n=324), microglia load correlated with p-tau load (rho=0.393, *p*<0.001).

To evaluate the effect of age at death, duration of illness, PSP stage, LBD stage, and ADNC level, ANCOVA test showed significant (**Figure 2A**) differences in the globus pallidus between groups 1 and 2 and 1 and 3 (group 1 the highest), in the putamen (1 and 2 > 3), furthermore, in the internal capsule, dorsomedial nucleus of the thalamus and subthalamic nucleus (1 > 3).

**Figure 2.**
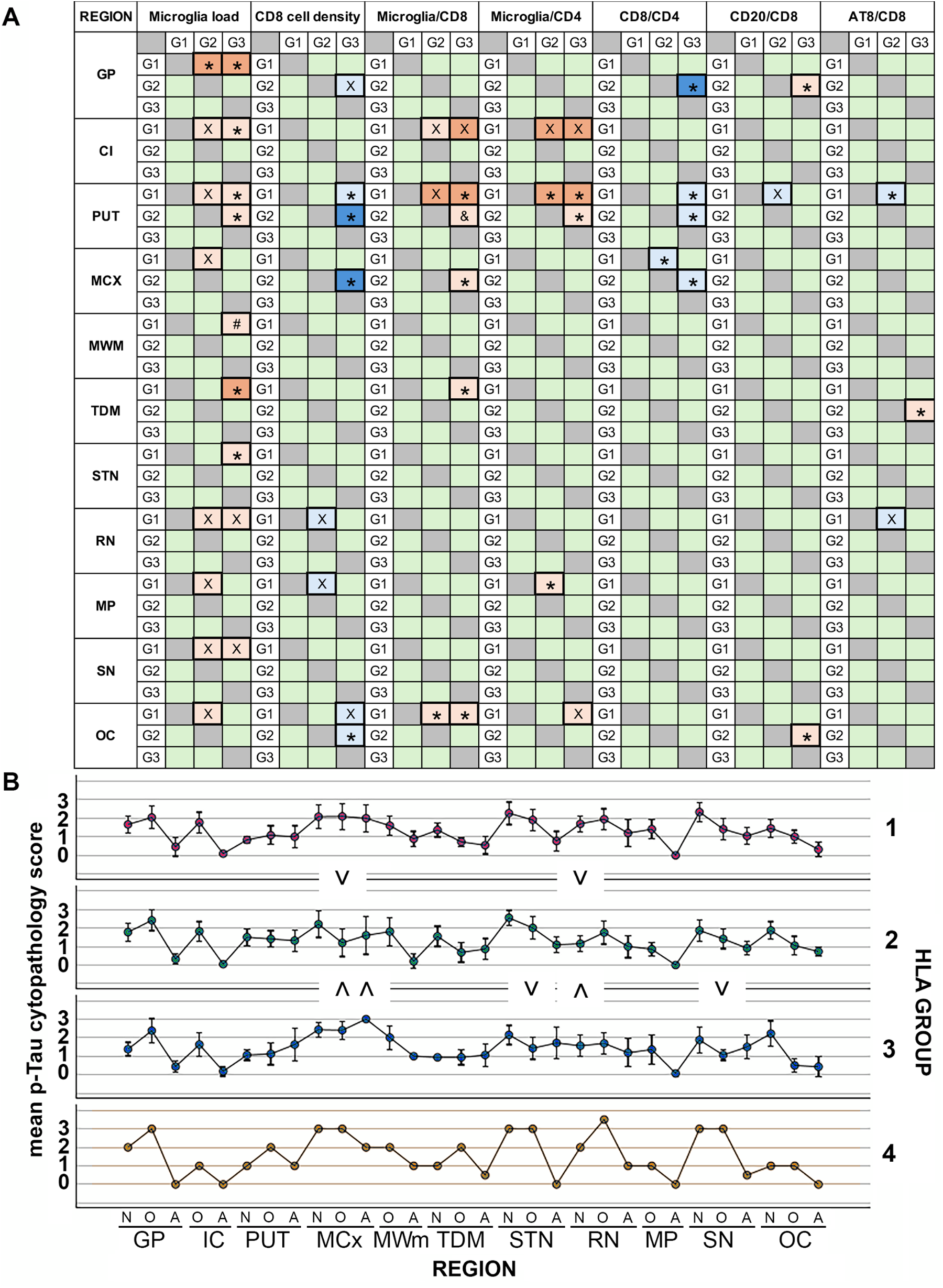
Summary of ANCOVA test results. A. Comparison of haplotype groups (G1, 2, 3) in anatomical regions following correction for age at death, PSP stage and ADNC level. Orange color indicates that the group in the horizontal row shows a higher value for the representative variable (dark orange color indicates *p*<0.01, light orange color indicates *p*<0.05). Blue color indicates that the group in the vertical columns shows a higher value for the representative variable (dark blue color indicates *p*<0.01, light blue color indicates *p*<0.05). X indicates significant only when corrected for age at death, PSP stage, ADNC but not duration of illness and not for stage of Lewy body pathology; *indicates significant when corrected for age, PSP, ADNC, duration of illness and for stage of Lewy body pathology; & indicates significant following correction for age, duration, ADNC, PSP and Lewy body pathology stage but not for correction for age, PSP, ADNC only; #indicates significant following correction for age, duration, ADNC, PSP but not after correction for Lewy body pathology stage. B. Error bars of p-tau cytopathology scores in different brain regions demonstrated for distinct HLA groups. Significant differences showing the higher values are indicated by a “V” and a turned “V”. Abbreviations: GP, globus pallidus; CI, capsula interna; PUT, putamen; MCX, motor cortex; MWM, motor white matter; TDM, thalamus dorsomedial nucleus; STN, subthalamic nucleus; RN, red nucleus; MP, midbrain peduncles; SN, substantia nigra; OC, oculomotor complex. N: neuronal; O: oligodendrocytic; A: astrocytic.

The anatomical lesion profile of microglia loads in different HLA haplotype groups (PSP stage ≥ 3) showed different patterns (**Figure 3A**). The midbrain and subthalamic nucleus were generally the highest, except for group 2 where the globus pallidus showed greater involvement. In other regions, groups 1 and 2 and the single case in group 4 showed similar patterns, and group 3 showed the most deviation (**online supplemental file Table S3**). The LNT case^21^ in group 2 showed higher p-tau and microglia load in several brain regions while the case with suspected postencephalitic parkinsonism^20^ showed lower microglia load and p-tau load in all regions, except the substantia nigra where p-tau load was the highest (**online supplemental file Figure S4**).

**Figure 3.**
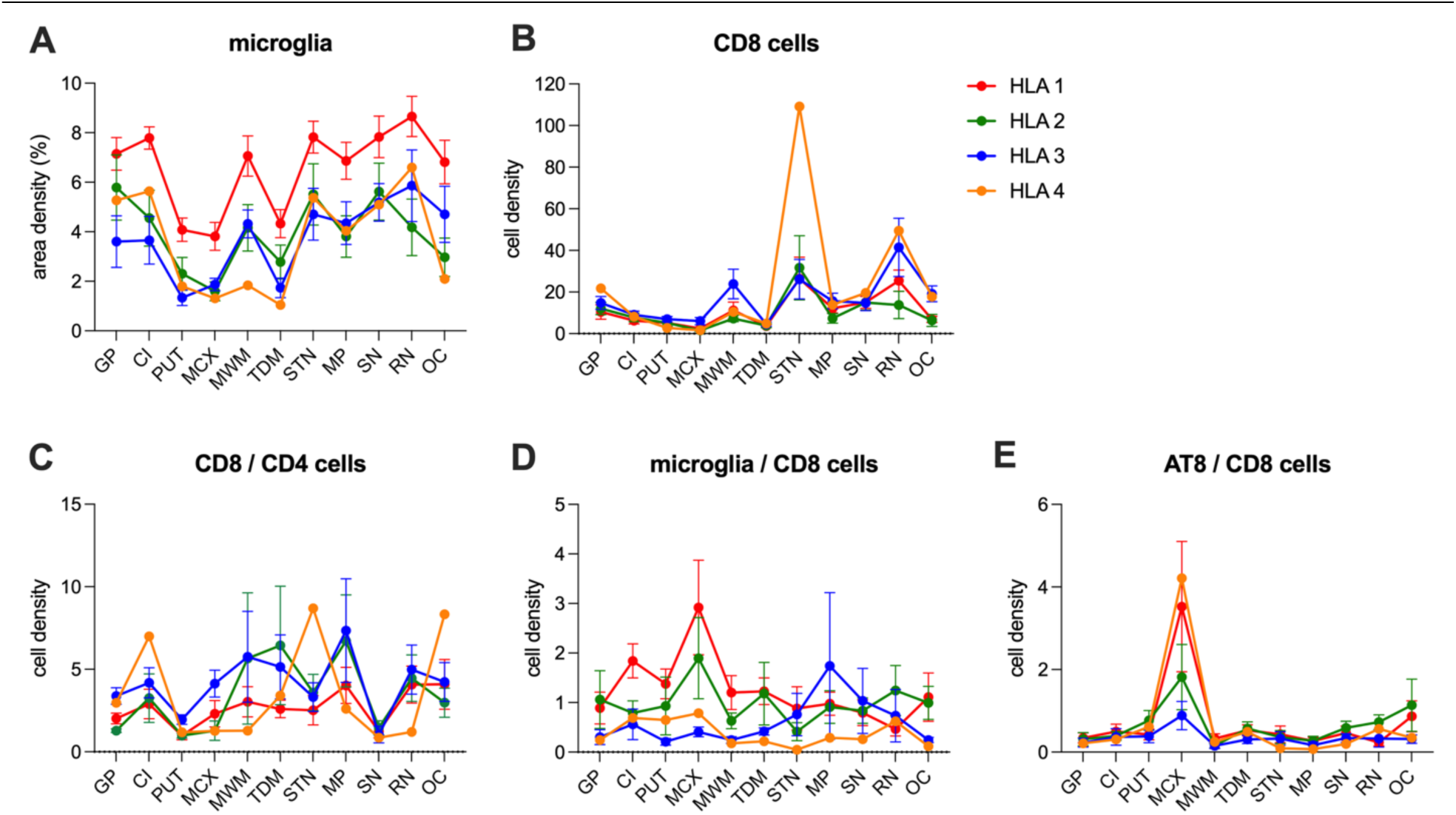
Microglia and inflammatory cell load and their ratios across anatomical regions in the different HLA haplotypes. Graphs show the microglia (A) and CD8 cell (B) load expressed as area and cell density, respectively, and the ratio of CD8/CD4 cells (C), microglia/CD8 cells (D) and AT8/CD8 cells (E) across brain regions expressed as the cell density in PSP > stage 2 cases. HLA haplotype groups are shown in different colours and show different patterns across brain regions: group 1 = red, group 2 = green, group 3 = blue and group 4 = yellow. Abbreviations: GP, globus pallidus; CI, capsula interna; PUT, putamen; MCX, motor cortex; MWM, motor white matter; TDM, thalamus dorsomedial nucleus; STN, subthalamic nucleus; RN, red nucleus; MP, midbrain peduncles; SN, substantia nigra; OC, oculomotor complex.

### Differences in CD8 cytotoxic cell counts between HLA haplotype groups

CD20 positive B cells and CD4 positive T cell densities were similar between groups. CD3 positive T cell densities were higher in the motor area white matter in group 3 compared to group 2. CD8 positive T cell densities were higher in group 3 versus group 1 and group 2 in several regions (**online Supplementary file Results**).

Pooling all regions and cases examined (n=324), densities of all cell types (CD20, CD3, CD4, and CD8) correlated with p-tau load (rho=0.114, 0.385, 0.189, 0352, respectively; *p*=0.040 for CD20 and *p*<0.001 for CD3, CD4 and CD8).

ANCOVA (determining the effect of age at death, duration of illness, PSP and LBD stage and ADNC level) test showed significant differences only for CD8 cytotoxic T cells in the putamen (group 3 > 1 and 3 > 2) motor cortex (3 > 2), and oculomotor complex (3 > 2) (**Figure 2A**).

The anatomical lesion profile of T cell types (e.g. CD8 cells; **Figure 3B**), and B cell density, in different HLA haplotype groups (PSP stage ≥ 3) showed different patterns. The subthalamic nucleus was amongst the most involved brain region examined with the presence of CD8 T cells and both the motor cortex and putamen the least involved. CD4 T cells were mostly present in the substantia nigra and subthalamic nucleus. As seen for microglia load, the globus pallidus was more affected by T cells in group 2 compared to other groups (**online supplemental file Table S3**). The LNT case^21^ in group 2 showed similar CD4 and CD8 cell density patterns and the case with suspected postencephalitic parkinsonism^20^ showed lower density patterns of CD4 and CD8 cells (**online supplemental file Figure S4**).

### Predictability of HLA haplotypes based on inflammatory marker-based pathology

Unsupervised GMM was used to evaluate whether natural clustering can occur based on the raw features of inflammatory-based pathology. This approach found no meaningful clustering and the unsupervised GMM resulted in low clustering accuracy (ARI = 0.0761 and clustering accuracy = 56.52%). Clustering performance significantly improved when RFC-extracted features were used for unsupervised clustering (ARI = 0.6671 and clustering accuracy = 86.96%). On the other hand, the predictability of HLA haplotypes based on the raw features of inflammatory-based pathology was low (prediction accuracy = 42.90%).

### Differences in total p-tau load and cytopathologies between HLA haplotype groups

Comparison of total p-tau load in the brain regions examined was similar between HLA haplotype groups (*p*>0.2 in all comparisons even following correction for age at death, duration of illness, PSP stage and ADNC). P-tau cytopathology scores were then compared between groups in cases with PSP stage ≥ 3. Mean neuronal, oligodendrocytic, and astrocytic scores showed variability between brain regions. Following correction for age at death, duration of illness, PSP stage and ADNC, ANCOVA test confirmed significant differences of p-tau cytopathology scores for oligodendrocytic (groups 1 > 2 and 3 > 2) and astrocytic (group 3 > 2) in the motor cortex, oligodendrocytic in the subthalamic nucleus (group 2 > 3) and substantia nigra (group 2 > 3), and neuronal in the red nucleus (group 1 > 2 and 3 > 2) (**Figure 2B**).

Based on the raw features of p-tau cytopathology, the unsupervised GMM had no meaningful clustering and low clustering accuracy (ARI = -0.0726 and clustering accuracy = 40.91%, respectively). Transformation of raw features into high-dimensional embedding space, clustering performance was improved (ARI = 0.1960 and clustering accuracy = 63.64%). The predictability of HLA haplotypes based on the raw features of p-tau cytopathology was low (prediction accuracy = 42.86%). Regional distribution patterns of total p-tau load varied between cases with PSP stage ≥ 3, exemplified by the more severe involvement of the motor cortex in group 3 and the single group 4 case compared to groups 1 and 2. The globus pallidus was more involved in group 2 than other groups (**online supplemental file Table S3**).

### Differences in ratios of neuropathological variables between HLA haplotype groups

Comparison of ratios of neuropathological variables showed significant variability between brain regions (**online Supplementary file Results**). Following correction for age at death duration of illness, PSP stage and ADNC level, ANCOVA test confirmed significant differences (*p*<0.05) (**Figure 2A**) for microglia/CD8 in the putamen (groups 1 and 2 > 3), motor cortex (2 > 3) and dorsomedial nucleus of the thalamus (1 > 3) and oculomotor complex (1 > 2 and 1> 3); microglia/CD4 in the putamen (1 > 2 and 1 >3 and 2 > 3) and midbrain peduncle (1 > 2); CD8/CD4 in the globus pallidus (3 > 2), putamen (3 > 1 and 3 > 2), and motor cortex (3 > 2 and 2 > 1); CD20/CD8 in the globus pallidus and oculomotor complex (2 > 3); AT8/CD8 in the putamen (2 > 1) and dorsomedial nucleus of the thalamus (2 > 3).

The anatomical lesion profile of ratios of neuropathological variables in different HLA haplotype groups (PSP stage >2) showed different patterns (**Figure 3 C-E**). Indeed, greater variability was observed between groups than observed for single variables (**online supplemental file Table S4**). The LNT case^21^ in group 2 showed similar regional distribution of ratios except for microglia/CD8, microglia/CD4 and CD8/CD4 differed in some anatomical regions (**online supplemental file Figure S5**). The case with suspected postencephalitic parkinsonism^20^ showed similar patterns except for the CD20/CD8 value, which deviated from other case in group 3 in some regions (**online supplemental file Figure S5**).

### Predictability of HLA haplotypes based on ratios of neuropathological variables

Unsupervised GMM was used to analyze the ratios of the inflammatory marker-based pathology to evaluate whether they naturally group into HLA haplotypes. This initially resulted in poor clustering and low clustering accuracy (ARI = 0.0278 and clustering accuracy = 52.17%), however, using the RFC-extracted features for unsupervised clustering significantly improved the clustering performance (ARI = 0.7446 and clustering accuracy = 91.30%; **Figure 4A**). Supervised RFC also resulted in a good prediction accuracy (prediction accuracy = 71.43%). Feature extraction revealed that the ratios between microglia and CD8, microglia and CD4, and CD8 and CD4 were the top features contributing to the classification of HLA haplotypes (**Figure 4B**).

**Figure 4.**
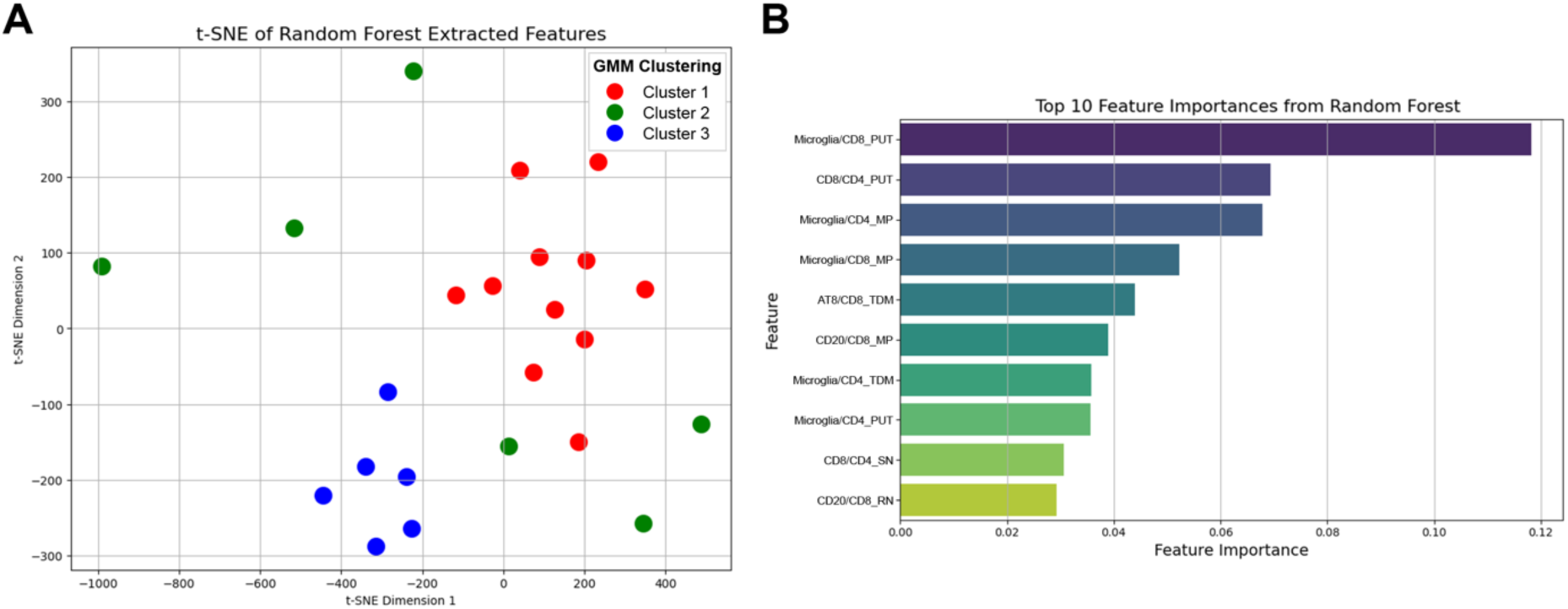
Visualization of the clusters generated by the unsupervised GMM based on the RFC-extracted features of ratios between inflammatory marker-based pathology. (**A**) t-SNE was used to reduce the high-dimensional data into two dimensions (component 1 and 2) and visualize the clusters of the ratios between inflammatory marker-based pathology from different regions using RFC-extracted features, where each datapoint represents a case; (**B**) the top 10 important features extracted from supervised RFC that predict the HLA haplotypes based on the raw features of ratios between inflammatory marker-based pathology. Red represents cluster 1, green represents cluster 2, and blue represents cluster 3 in panel **A**, longer bar graphs represent higher importance in distinguishing the HLA haplotypes in panel **B**. It is important to note that t-SNE reduces dimensionality and therefore, visualization may not represent true clustering structures. Abbreviations: CI, capsula interna; GMM, Gaussian Mixture Model; GP, globus pallidus; MP, midbrain peduncles; OC, oculomotor complex; PUT, putamen; RN, red nucleus; SN, substantia nigra; TDM, thalamus dorsomedial nucleus; t-SNE, t-distributed Stochastic Neighbor Embedding.

## Discussion

Observations from this study open a novel conceptual approach in the interpretation of inflammatory responses in the human brain affected by PSP with implications for other neurodegenerative diseases. Indeed, we show that the distribution and combination of microglia, T and B cells are strongly associated with the HLA haplotype/allele an individual with PSP carries. This would also suggest that the common neuropathology described as PSP-type could have different initiating etiopathogenic events or inflammatory components that contribute to progression. Analogously the finding of the same tau filament fold in chronic traumatic encephalopathy, the measles virus-related subacute sclerosing panencephalitis,^22^ and amyotrophic lateral sclerosis/parkinsonism-dementia complex,^23^ interpreted as environmentally induced in addition to the dominantly inherited mutation D395G in the gene encoding valosin-containing protein causing vacuolar tauopathy,^24^ indicates that common neuropathology and tau filament fold can be induced by various etiologies. Our recent study already raised the likelihood that some PSP cases might have an autoimmune mechanism.^15^ This was supported by finding CD8 cytotoxic T cell-neuron contacts in the substantia nigra in some cases^11^ and here by showing very different patterns of components interpreted in the frame of the so called neuroinflammatory response.^6^

In our cohort, cases with the haplotype described in narcolepsy (*DRB1*\*15:01-*DQB1*\*06:02)^25^ showed a greater microglia reaction compared to those with other haplotypes. In contrast, CD8 T cell response was different in other haplotypes reminiscent of that observed in the single case with the haplotype frequent in IgLON5-associated encephalitis (*DRB1*\*10:01-*DQB1*\*05:01).^7, 26^ The relevance of the HLA locus in PSP is strongly supported by genome-wide association studies of PSP indicating the presence of risk factor(s) at 6p21^27^ including genes in the vicinity to the *HLA* locus (e.g., *TNXB*).^28^ Interestingly, the LNT case^21^ showed greater microglia activation than other cases in group 2 but generally showed similar patterns, while the case with suspected postencephalitic parkinsonism and longest duration of illness showed less inflammatory pathology, except for some regions with increased CD20 positive B cells over CD8 positive cytotoxic T cells. Altogether, this suggests that rare subtypes can differ beyond the effect of HLA haplotype and might have individual patterns.

The role of cellular adaptive immunity in PSP was discussed in a study focusing on the frontal cortex of cases with clinical frontotemporal dementia associated with PSP pathology.^29^ Studies focusing on peripheral markers support this notion, but also suggest variability between patients,^12,13^ as seen in our present study. The differences in patterns of inflammatory pathology remained significant in PSP strategic regions following the correction for neurodegenerative co-pathologies that could have an impact of neuroinflammation. Importantly, recent experimental models suggest the significance of T cells in the progression of tauopathy^30^ raising the option of T cells as a prime therapy target instead of neuronal tangles. Correlation of CD8 T cells with p-tau pathology was also discussed in human brain studies,^29^ as seen in our study.

Although the clinical symptoms alone might not be helpful to predict the HLA haplotype, the patterns of the sequences of clinical symptoms with duration of illness showed a relatively high prediction accuracy of (80%). It must be noted that one limitation of this study is that some cases included only retrospective clinical information and there was a lack of disease scores that would have allowed even better prediction. The unbiased evaluation of the patterns can be interpreted as follows. First, when the raw features were used for unsupervised clustering, the cases did not naturally cluster into the different HLA haplotypes, likely due to noise in the dataset (i.e., irrelevant regions that do not have differences across the features). However, when RFC-extracted features that transformed the raw features into a high-dimensional embedding space were used for unsupervised clustering as a second level of analysis, clustering performance increased. This suggests that the relationship between the HLA haplotypes and these features are non-linear and complex, but HLA haplotypes are still distinguishable with high accuracy, supporting the notion that the overall patterns of pathology variables differ in HLA haplotypes. This is supported by the predictability of some constellation of variables that were helpful in predicting HLA haplotypes. It is important to note that high prediction accuracy could be due to overfitting, therefore, further evaluation of larger number of cases could reveal stronger predictive power of these features.

In conclusion, our study emphasizes the importance of integrating HLA haplotyping into the interpretation of studies on neuroinflammation and related biomarkers. This conceptual approach might be relevant for Parkinson’s disease and AD, where HLA was examined but not correlated with markers of neuroinflammation.^31–33^ Since potential epitopes within the tau peptide, but not the misfolded tau filament, may bind to alleles overexpressed in PSP patients, the possibility that a subset of PSP cases might have an autoimmune pathophysiological component is suggested.^15^ Our findings provide further evidence advocating for subtyping and stratifying patients for disease modifying treatments and encourage the consideration of therapeutic targets beyond tau, particularly immune modulatory agents in a subset of patients. In addition, genome-wide association and biomarker studies could consider re-analyzing datasets stratified for HLA haplotype groups, which would prove more informative in deciphering the involved pathways.

## Data Availability

Data will be shared upon reasonable request from any qualified investigator.

## Acknowledgements

The authors especially acknowledge the patients and their families for their donation. GGK holds the Rossy Chair in PSP research at UHN.

## Author contributions

Concept and design of the study: SLF, GGK

Acquisition and analysis of data: SLF, SZ, AK, HT, HC, JL, SF, JW, MCT, AEL, GGK

Drafting a significant portion of the manuscript or figures: SLF, GGK

