## Supplementary material for "Neuroinflammation distinguishes HLA haplotypes in progressive supranuclear palsy": online supplemental file

### SUPPLEMENTARY METHODS

Scanned images of immunostained sections were imported into HALO software (version 3.6.4134, Indica Labs), and using the Pen Annotation tool, 11 subregions of interest were manually annotated: putamen, globus pallidus and capsula interna of the basal ganglia, gray matter and white matter of the motor cortex, medial nucleus and subthalamic nucleus of the thalamus, cerebral peduncle, substantia nigra, ruber and oculomotor complex of the midbrain (Fig. S1A). The Object Colocalization module (version 2.1.5) within HALO software was then used for the automated quantification of DAB-positive staining to determine the area densities of AT8 (Fig. S1B,C) and HLA-DR (Fig. S1D,E). Area density for AT8 and HLA-DR was expressed as the percentage of the DAB-positive area within the area of the subregion of interest. For automated counts of CD3, CD4, CD8 and CD20 cells in HALO, the detection parameters within the Object Colocalization module were initially adjusted such that only single immuno-positive cells were detected (Fig. S2A-B). Using these preliminary parameters, 14-15 randomly selected immunostained sections were analyzed and the average area of single CD3, CD4, CD8 and CD20 cells was calculated. The average area ($\mu m^2$±SEM, n=number of cells counted) calculated for each immuno-positive cell was as follows: CD3 (20.96±0.05, n=63,097), CD4 (20.70±0.08, n=14,439), CD8 (26.21±0.08, n=32,980), CD20 (19.94±0.12, n=8,265). Finally, the parameters were re-adjusted such that the total area of CD3, CD4, CD8 and CD20 immuno-positivity was detected within the subregions of interest (Fig. S2C), and then this total immuno-positive area was divided by the calculated average area of a single cell to give the total number of immuno-positive cells detected within the subregion. CD3, CD4, CD8 and CD20 area density is expressed as the number of DAB-positive cells per $mm^2$ of the subregion of interest. Detailed parameters utilized for the detection of immuno-positive area with the Object Colocalization module can be found in Tables S1 and S2. False immuno-positive detection of neuromelanin in the substantia nigra subregion was excluded in all analyses by manually screening and circling neuromelanin in the substantia nigra using the Exclusion Annotation tool in Halo (Fig. S3).

**Figure S1: Annotation of brain subregions of interest and automated detection of AT8 and HLA-DR area density using HALO software.** (A) Representative annotation of the ruber (purple outline), substantia nigra (yellow outline) and cerebral peduncle (green outline) in the midbrain using HALO. (B) Sample AT8 immunostaining and (C) AT8 immuno-positive area detection (green) in HALO. (D) Sample HLA-DR immunostaining and (E) HLA-DR immuno-positive area detection (green) in HALO. Scale bar = 1mm in (A), 20µm in (B-E).

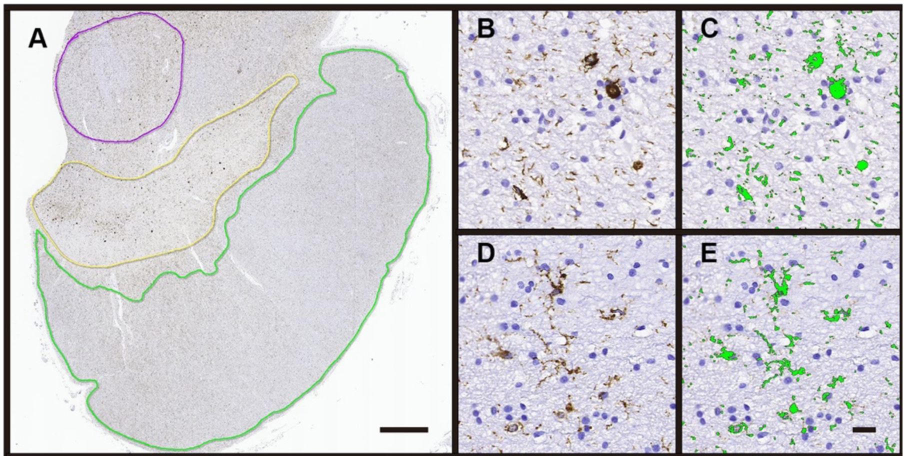

**Figure S2: Automated detection of CD3, CD4, CD8 and CD20 immunoreactivity and area density calculation using HALO software.** (A) Representative image of CD8 immunostaining in the capsula interna of the basal ganglia. (B) Detection parameters within the Object Colocalization module of HALO were initially adjusted such that only single immuno-positive cells were detected (green). This allowed the average area of a single immuno-positive (e.g. CD8) cell to be calculated. (C) Detection parameters were then re-adjusted so that the total immunoreactive area was detected (green). The total immunoreactive area was then divided by the average area of a single cell (calculated from B) to determine the total number of immuno-positive cells detected within the subregion of interest. Scale bar = 20μm.

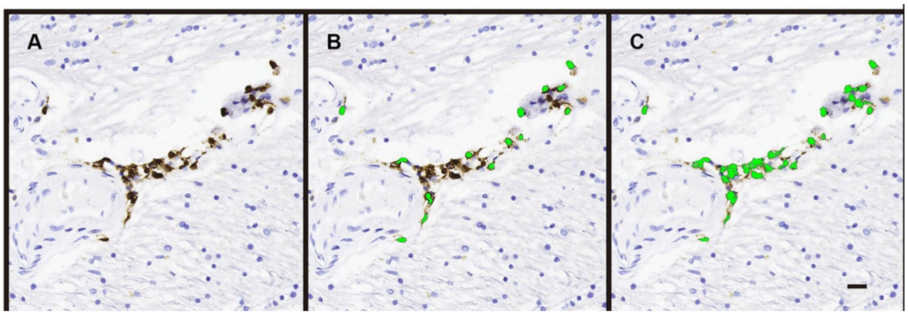


**Figure S3: Method of neuromelanin exclusion from analysis in the substantia nigra by HALO software.** A) Representative image of neuromelanin (black arrows) and AT8 immunostaining in the substantia nigra. (B) Regions of neuromelanin were manually circled (yellow/black dashed circles) and excluded from analysis using the Exclusion Annotation tool in HALO. C) AT8 immunostaining is detected (green) and neuromelanin has been excluded from analysis. Scale bar = 20μm.

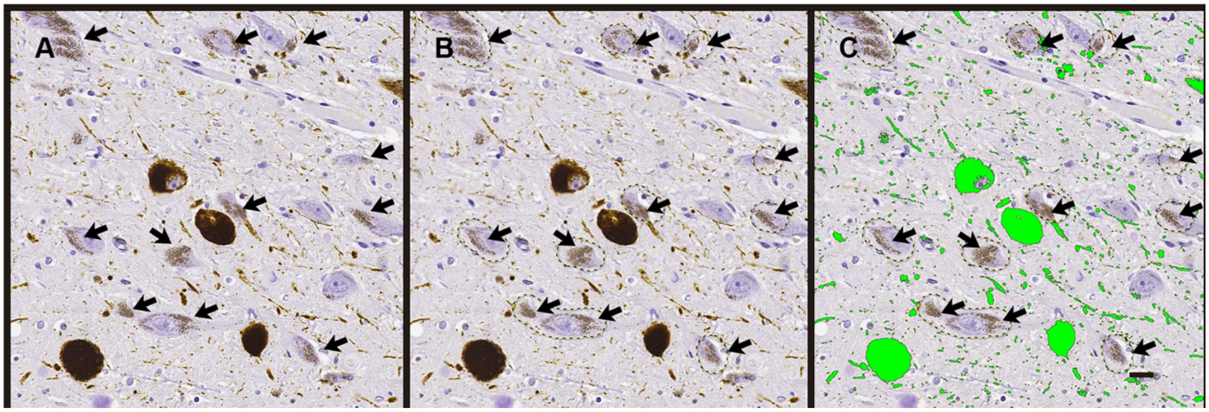

**Table S1:** Object Colocalization module parameters in HALO software for the automated detection of the total area of AT8, HLA-DR, CD3, CD4, CD8 and CD20 immunostaining.

| Parameters | AT8 | HLA-DR | CD3 | CD4 | CD8 | CD20 |
|---|---|---|---|---|---|---|
| **Stain Color** | 0.467, 0.600, 0.773 | 0.467, 0.600, 0.773 | 0.394, 0.467, 0.509 | 0.394, 0.467, 0.509 | 0.394, 0.467, 0.509 | 0.394, 0.467, 0.509 |
| **Blur Radius** | 0 | 0 | 0 | 0 | 0 | 0 |
| **Contrast Radius** | 278.444 | 500 | 500 | 500 | 500 | 500 |
| **Contrast Threshold** | 0.821 | 0.828 | 0.841 | 0.841 | 0.841 | 0.841 |
| **Optical Density** | 0 | 0 | 0.69 | 0.69 | 0.69 | 0.69 |
| **Object Size** | 10, 100000 | 10, 100000 | 15, 100000 | 15, 100000 | 15, 100000 | 15, 100000 |
| **Connect Length** | 0 | 0 | 0 | 0 | 0 | 0 |
| **Fill Holes** | False | False | True | True | True | True |

**Table S2:** Object Colocalization module parameters in HALO software for the automated detection of area in single CD3, CD4, CD8 and CD20 immuno-positive cells.

| Parameters | CD3 | CD4 | CD8 | CD20 |
|---|---|---|---|---|
| **Stain Color** | 0.394, 0.467, 0.509 | 0.394, 0.467, 0.509 | 0.394, 0.467, 0.509 | 0.394, 0.467, 0.509 |
| **Blur Radius** | 11.73 | 22.26 | 18.27 | 15.09 |
| **Contrast Radius** | 10 | 9 | 9 | 9 |
| **Contrast Threshold** | 0.868 | 0.861 | 0.861 | 0.861 |
| **Optical Density** | 0.47 | 0.47 | 0.47 | 0.47 |
| **Object Size** | 10, 100000 | 10, 100000 | 10, 100000 | 10, 100000 |
| **Connect Length** | 0 | 0 | 0 | 0 |
| **Fill Holes** | True | True | True | True |

### SUPPLEMENTARY RESULTS

#### *Microglia load: comparisons without corrections*

Microglia load was higher in the internal capsule (*p*=0.043), motor cortex (*p*=0.036), cerebral peduncle (*p*=0.040), substantia nigra (*p*=0.049), red nucleus and superior cerebellar peduncle (*p*=0.039), and oculomotor complex (*p*=0.011), between group 1 and 2 cases. Microglial load was higher in the globus pallidus (*p*=0.037), internal capsule (*p*=0.008), putamen (*p*=0.003), dorsomedial thalamus (*p*=0.005) and substantia nigra (*p*=0.016) in group 1 versus group 3 cases.

#### *CD20, CD3, CD4, and CD8 cytotoxic cell counts: comparisons without corrections*

CD20 positive B cells and CD4 positive T cell densities were similar between groups. CD3 positive T cell densities were higher in the motor area white matter in group 3 compared to group 2. In the motor cortex CD8 positive T cell densities were higher in group 3 versus group 1 (*p*=0.036) and group 2 (*p*=0.007), and in the white matter (*p*=0.036 for Groups 3 and 1; *p*=0.040 for group 3 versus 2). In the red nucleus/superior cerebellar peduncle CD8 positive T cell densities were higher in group 1 compared to group 2 (*p*=0.048). Finally, in the oculomotor complex group 3 showed higher CD8 positive T cell densities than group 2 (*p*=0.005).

#### *p-Tau cytopathologies: comparisons without corrections*

P-tau cytopathology scores were compared between groups in cases with PSP stage $\geq$ 3. Mean oligodendrocytic (*p*=0.001) and astrocytic (*p*=0.032) scores were higher in the motor cortex, neuronal scores higher in the red nucleus (*p*=0.029), and oligodendrocytic scores higher (*p*=0.012) in the cerebral peduncle in group 1 compared to group 2. The neuronal p-tau cytopathology score was higher in the putamen (*p*=0.016) in group 2. Mean neuronal scores were higher in the dorsomedial thalamic nucleus (*p*=0.003) and oligodendrocytic scores were higher in the oculomotor complex (*p*=0.018) in group 1 versus group 3 cases, while neuronal scores were higher in the oculomotor complex (*p*=0.003) and astrocytic scores higher in the motor cortex (*p*=0.039) in group 3 cases. Finally, oligodendrocytic (*p*=0.009) and astrocytic (*p*=0.016) p-tau cytopathology scores were higher in the motor cortex and neuronal scores higher in the oculomotor complex (*p*=0.034) in group 3 versus 2, while neuronal scores in the dorsomedial thalamic nucleus (*p*=0.02) and astrocytic p-tau scores in the oculomotor complex (*p*=0.002) were higher in group 2 versus 3 cases.

#### *Differences in ratios of neuropathological variables: comparisons without corrections*

Comparison of ratios of neuropathological variables revealed higher values in group 1 compared to 3 in the putamen and internal capsule for microglia/CD8 (*p*=0.003 and 0.004, respectively) and microglia/CD4 (*p*=0.003 and 0.026, respectively); furthermore in the motor cortex (*p*=0.037), motor area white matter (*p*=0.008), and dorsomedial nucleus of the thalamus (*p*=0.014), and oculomotor nucleus (*p*=0.024) for microglia/CD8. Higher values were observed in group 1 compared to 2 in the putamen and internal capsule for microglia/CD8 (*p*=0.014 and 0.041, respectively) and microglia/CD4 (*p*=0.005 and p=0.022, respectively). CD20/CD8 in the putamen (*p*=0.045) and AT8/CD8 in the red nucleus/superior cerebellar peduncle (*p*=0.026) were higher in group 2 compared to 1. CD8/CD4 was higher in group 3 than 2 in the putamen (*p*=0.031), globus pallidus (*p*=0.008), and motor cortex (p=0.012); furthermore, in the putamen between group 3 and 1 (3 > 1; *p*=0.030). Finally, microglia/CD8 was higher in the motor cortex in group 2 versus 3 (*p*=0.031) and oculomotor complex (*p*=0.042).

**Table S3**. Descriptive statistics of neuropathological variables in different brain regions in distinct groups of cases (all PSP stage > 2) defined based on HLA haplotypes (see manuscript). Color coding indicates the different brain regions. Abbreviations: GP, globus pallidus; CI, capsula interna; PUT, putamen; MCX, motor cortex; MWM, motor white matter; TDM, thalamus dorsomedial nucleus; STN, subthalamic nucleus; RN, red nucleus; MP, midbrain peduncles; SN, substantia nigra; OC, oculomotor complex.

| | | GROUP 1 | | | | | GROUP 2 | | | | | GROUP 3 | | | | | GROUP 4 |
|---|---|---|---|---|---|---|---|---|---|---|---|---|---|---|---|---|---|---|
| | | N | Mean | SD | SE | | N | Mean | SD | SE | | N | Mean | SD | SE | | N | Mean |
| AT8 | OC | 9 | 5.29 | 1.58 | 0.53 | STN | 9 | 5.92 | 3.95 | 1.32 | OC | 7 | 5.72 | 3.93 | 1.48 | STN | 1 | 10.17 |
| | STN | 7 | 5.09 | 3.87 | 1.46 | SN | 9 | 4.46 | 2.77 | 0.92 | STN | 6 | 5.70 | 3.05 | 1.24 | MCX | 1 | 7.00 |
| | RN | 9 | 4.39 | 2.34 | 0.78 | GP | 7 | 4.02 | 3.86 | 1.46 | MCX | 5 | 3.78 | 1.12 | 0.50 | RN | 1 | 6.30 |
| | SN | 9 | 3.84 | 1.01 | 0.34 | OC | 8 | 3.66 | 3.34 | 1.18 | RN | 7 | 3.76 | 2.40 | 0.91 | OC | 1 | 6.13 |
| | MCX | 8 | 3.62 | 2.03 | 0.72 | MCX | 6 | 3.34 | 3.83 | 1.56 | SN | 7 | 3.17 | 1.71 | 0.65 | GP | 1 | 4.59 |
| | GP | 9 | 2.41 | 1.71 | 0.57 | RN | 9 | 3.07 | 2.01 | 0.67 | GP | 6 | 2.44 | 2.01 | 0.82 | SN | 1 | 3.91 |
| | CI | 9 | 2.23 | 1.70 | 0.57 | TDM | 9 | 2.47 | 2.39 | 0.80 | CI | 6 | 2.28 | 1.81 | 0.74 | MWM | 1 | 2.57 |
| | MWM | 8 | 2.08 | 1.79 | 0.63 | PUT | 9 | 2.40 | 1.81 | 0.60 | MWM | 5 | 2.23 | 0.86 | 0.38 | CI | 1 | 2.53 |
| | MP | 9 | 1.66 | 0.67 | 0.22 | CI | 9 | 2.15 | 2.05 | 0.68 | PUT | 7 | 1.69 | 1.61 | 0.61 | TDM | 1 | 2.37 |
| | TDM | 9 | 1.61 | 0.82 | 0.27 | MWM | 6 | 1.61 | 1.45 | 0.59 | MP | 7 | 1.56 | 1.63 | 0.62 | PUT | 1 | 1.70 |
| | PUT | 9 | 1.26 | 1.25 | 0.42 | MP | 9 | 0.95 | 0.59 | 0.20 | TDM | 7 | 1.34 | 1.02 | 0.39 | MP | 1 | 0.99 |
| MG | RN | 9 | 8.66 | 2.44 | 0.81 | GP | 7 | 5.80 | 3.50 | 1.32 | RN | 7 | 5.86 | 3.83 | 1.45 | RN | 1 | 6.59 |
| | SN | 9 | 7.83 | 2.51 | 0.84 | SN | 9 | 5.62 | 3.45 | 1.15 | SN | 7 | 5.19 | 2.00 | 0.76 | CI | 1 | 5.64 |
| | STN | 8 | 7.82 | 1.83 | 0.65 | STN | 9 | 5.51 | 3.70 | 1.23 | STN | 6 | 4.71 | 2.56 | 1.05 | STN | 1 | 5.37 |
| | CI | 9 | 7.79 | 1.35 | 0.45 | CI | 9 | 4.55 | 3.39 | 1.13 | OC | 7 | 4.71 | 2.99 | 1.13 | GP | 1 | 5.27 |
| | GP | 9 | 7.15 | 1.97 | 0.66 | RN | 9 | 4.18 | 3.42 | 1.14 | MP | 7 | 4.35 | 2.28 | 0.86 | SN | 1 | 5.09 |
| | MWM | 8 | 7.06 | 2.30 | 0.81 | MWM | 6 | 4.16 | 2.30 | 0.94 | MWM | 5 | 4.32 | 1.26 | 0.56 | MP | 1 | 4.04 |
| | MP | 9 | 6.86 | 2.23 | 0.74 | MP | 9 | 3.81 | 2.51 | 0.84 | CI | 6 | 3.66 | 2.36 | 0.96 | OC | 1 | 2.09 |
| | OC | 9 | 6.82 | 2.64 | 0.88 | OC | 8 | 2.97 | 2.19 | 0.77 | GP | 6 | 3.60 | 2.55 | 1.04 | MWM | 1 | 1.84 |
| | TDM | 9 | 4.33 | 1.70 | 0.57 | TDM | 9 | 2.78 | 2.05 | 0.68 | MCX | 5 | 1.87 | 0.56 | 0.25 | PUT | 1 | 1.80 |
| | PUT | 9 | 4.08 | 1.41 | 0.47 | PUT | 9 | 2.30 | 1.99 | 0.66 | TDM | 7 | 1.74 | 1.05 | 0.40 | MCX | 1 | 1.31 |
| | MCX | 8 | 3.81 | 1.59 | 0.56 | MCX | 6 | 1.59 | 0.94 | 0.38 | PUT | 7 | 1.33 | 0.83 | 0.31 | TDM | 1 | 1.05 |
| CD3 | SN | 9 | 19.82 | 12.16 | 4.05 | SN | 9 | 21.34 | 14.62 | 4.87 | RN | 7 | 30.84 | 29.99 | 11.34 | STN | 1 | 32.42 |
| | RN | 9 | 19.34 | 11.39 | 3.80 | GP | 7 | 14.50 | 8.60 | 3.25 | SN | 7 | 23.65 | 7.26 | 2.74 | RN | 1 | 20.25 |
| | STN | 8 | 19.08 | 22.13 | 7.82 | STN | 9 | 12.65 | 14.58 | 4.86 | MWM | 5 | 16.48 | 9.15 | 4.09 | SN | 1 | 15.69 |
| | GP | 9 | 12.01 | 12.35 | 4.12 | RN | 9 | 10.75 | 13.70 | 4.57 | STN | 6 | 15.61 | 12.07 | 4.93 | GP | 1 | 11.36 |
| | MWM | 8 | 7.66 | 6.47 | 2.29 | PUT | 9 | 9.76 | 3.97 | 1.32 | GP | 6 | 11.59 | 8.89 | 3.63 | MP | 1 | 9.25 |
| | MP | 9 | 7.06 | 4.50 | 1.50 | OC | 8 | 5.40 | 4.85 | 1.72 | OC | 7 | 11.14 | 7.82 | 2.95 | TDM | 1 | 8.66 |
| | OC | 9 | 6.42 | 3.65 | 1.22 | MWM | 6 | 4.96 | 4.06 | 1.66 | MP | 7 | 8.55 | 6.27 | 2.37 | MWM | 1 | 8.37 |
| | PUT | 9 | 6.42 | 4.89 | 1.63 | CI | 9 | 4.27 | 3.83 | 1.28 | PUT | 7 | 7.64 | 2.72 | 1.03 | OC | 1 | 4.25 |
| | CI | 9 | 4.91 | 2.58 | 0.86 | MP | 9 | 3.27 | 3.91 | 1.30 | CI | 6 | 6.28 | 3.75 | 1.53 | CI | 1 | 3.53 |
| | MCX | 8 | 4.57 | 5.18 | 1.83 | TDM | 9 | 3.15 | 3.38 | 1.13 | MCX | 5 | 3.39 | 0.56 | 0.25 | PUT | 1 | 3.06 |
| | TDM | 9 | 3.06 | 2.73 | 0.91 | MCX | 6 | 2.58 | 1.72 | 0.70 | TDM | 7 | 3.06 | 1.89 | 0.72 | MCX | 1 | 1.29 |
| CD4 | SN | 9 | 21.47 | 15.69 | 5.23 | SN | 9 | 18.03 | 18.63 | 6.21 | SN | 7 | 20.00 | 10.90 | 4.12 | SN | 1 | 22.76 |
| | STN | 8 | 9.83 | 8.74 | 3.09 | GP | 7 | 11.53 | 9.88 | 3.73 | RN | 7 | 8.65 | 9.17 | 3.47 | STN | 1 | 12.55 |
| | RN | 9 | 8.33 | 6.04 | 2.01 | STN | 9 | 10.90 | 13.35 | 4.45 | OC | 7 | 6.89 | 6.81 | 2.57 | MWM | 1 | 8.18 |
| | GP | 9 | 6.15 | 3.32 | 1.11 | PUT | 9 | 7.44 | 5.69 | 1.90 | MWM | 5 | 6.88 | 5.97 | 2.67 | GP | 1 | 7.32 |
| | PUT | 9 | 5.40 | 2.77 | 0.92 | TDM | 9 | 6.24 | 9.89 | 3.30 | STN | 6 | 6.51 | 5.52 | 2.25 | RN | 1 | 7.26 |
| | OC | 9 | 5.09 | 4.76 | 1.59 | RN | 9 | 6.17 | 5.99 | 2.00 | GP | 6 | 4.76 | 2.97 | 1.21 | MP | 1 | 5.31 |
| | MWM | 8 | 4.09 | 2.47 | 0.87 | CI | 9 | 5.59 | 5.44 | 1.81 | PUT | 7 | 3.96 | 2.01 | 0.76 | PUT | 1 | 2.35 |
| | MP | 9 | 3.68 | 2.14 | 0.71 | OC | 8 | 5.58 | 10.06 | 3.56 | MP | 7 | 3.35 | 2.57 | 0.97 | OC | 1 | 2.13 |
| | CI | 9 | 3.34 | 2.43 | 0.81 | MWM | 6 | 5.02 | 3.56 | 1.45 | CI | 6 | 2.36 | 1.40 | 0.57 | TDM | 1 | 1.41 |
| | TDM | 9 | 3.10 | 2.48 | 0.83 | MP | 9 | 2.77 | 3.05 | 1.02 | TDM | 7 | 2.23 | 2.58 | 0.98 | MCX | 1 | 1.32 |
| | MCX | 8 | 1.77 | 1.04 | 0.37 | MCX | 6 | 2.66 | 2.38 | 0.97 | MCX | 5 | 1.15 | 0.75 | 0.33 | CI | 1 | 1.17 |
| CD8 | STN | 8 | 26.48 | 28.97 | 10.24 | STN | 9 | 31.65 | 46.06 | 15.35 | RN | 7 | 41.46 | 37.10 | 14.02 | STN | 1 | 109.13 |
| | RN | 9 | 25.46 | 15.30 | 5.10 | SN | 9 | 14.98 | 10.96 | 3.65 | STN | 6 | 26.16 | 23.30 | 9.51 | RN | 1 | 49.43 |
| | SN | 9 | 15.01 | 8.53 | 2.84 | RN | 9 | 13.77 | 19.66 | 6.55 | MWM | 5 | 23.83 | 15.87 | 7.10 | GP | 1 | 21.73 |
| | MP | 9 | 12.16 | 6.75 | 2.25 | GP | 7 | 11.45 | 8.49 | 3.21 | OC | 7 | 19.16 | 10.10 | 3.82 | SN | 1 | 19.56 |
| | MWM | 8 | 11.13 | 11.30 | 4.00 | CI | 9 | 7.80 | 6.26 | 2.09 | MP | 7 | 15.69 | 9.87 | 3.73 | OC | 1 | 17.71 |
| | GP | 9 | 10.41 | 10.28 | 3.43 | MP | 9 | 7.39 | 6.88 | 2.29 | GP | 6 | 14.73 | 7.53 | 3.07 | MP | 1 | 13.87 |
| | CI | 9 | 6.25 | 4.80 | 1.60 | MWM | 6 | 7.28 | 3.69 | 1.51 | SN | 7 | 14.73 | 9.70 | 3.67 | MWM | 1 | 10.45 |
| | OC | 9 | 6.10 | 4.50 | 1.50 | OC | 8 | 6.34 | 8.10 | 2.86 | CI | 6 | 9.04 | 4.52 | 1.84 | CI | 1 | 8.17 |
| | PUT | 9 | 5.12 | 5.63 | 1.88 | PUT | 9 | 5.13 | 4.45 | 1.48 | PUT | 7 | 6.93 | 3.47 | 1.31 | TDM | 1 | 4.82 |
| | TDM | 9 | 3.62 | 2.36 | 0.79 | TDM | 9 | 3.82 | 2.84 | 0.95 | MCX | 5 | 6.04 | 3.93 | 1.76 | PUT | 1 | 2.77 |
| | MCX | 8 | 2.41 | 1.98 | 0.70 | MCX | 6 | 1.53 | 1.07 | 0.44 | TDM | 7 | 4.55 | 2.77 | 1.05 | MCX | 1 | 1.66 |
| CD20 | SN | 9 | 3.04 | 2.78 | 0.93 | GP | 7 | 3.16 | 2.28 | 0.86 | SN | 7 | 2.98 | 1.61 | 0.61 | SN | 1 | 1.58 |
| | GP | 9 | 1.23 | 1.24 | 0.41 | PUT | 9 | 2.29 | 1.87 | 0.62 | PUT | 7 | 1.93 | 1.70 | 0.64 | RN | 1 | 1.29 |
| | STN | 8 | 1.16 | 2.23 | 0.79 | SN | 9 | 2.11 | 1.35 | 0.45 | GP | 6 | 1.44 | 0.83 | 0.34 | MP | 1 | 0.60 |
| | PUT | 9 | 1.10 | 0.79 | 0.26 | CI | 9 | 1.25 | 1.69 | 0.56 | RN | 7 | 0.92 | 1.53 | 0.58 | MWM | 1 | 0.59 |
| | RN | 9 | 0.80 | 1.27 | 0.42 | OC | 8 | 0.51 | 0.89 | 0.32 | MWM | 5 | 0.91 | 1.20 | 0.54 | GP | 1 | 0.46 |
| | MWM | 8 | 0.61 | 0.48 | 0.17 | STN | 9 | 0.46 | 0.40 | 0.13 | OC | 7 | 0.59 | 0.73 | 0.28 | TDM | 1 | 0.24 |
| | MCX | 8 | 0.40 | 0.28 | 0.10 | MWM | 6 | 0.46 | 0.41 | 0.17 | MCX | 5 | 0.57 | 0.58 | 0.26 | MCX | 1 | 0.22 |
| | MP | 9 | 0.33 | 0.22 | 0.07 | MCX | 6 | 0.32 | 0.30 | 0.12 | MP | 7 | 0.52 | 0.54 | 0.21 | PUT | 1 | 0.19 |
| | OC | 9 | 0.32 | 0.48 | 0.16 | MP | 9 | 0.23 | 0.23 | 0.08 | STN | 6 | 0.42 | 0.38 | 0.15 | CI | 1 | 0.11 |
| | TDM | 9 | 0.17 | 0.12 | 0.04 | TDM | 9 | 0.21 | 0.14 | 0.05 | TDM | 7 | 0.34 | 0.46 | 0.17 | STN | 1 | 0.00 |
| | CI | 9 | 0.16 | 0.16 | 0.05 | RN | 9 | 0.18 | 0.12 | 0.04 | CI | 6 | 0.33 | 0.29 | 0.12 | OC | 1 | 0.00 |

**Table S4**. Descriptive statistics of ratios of neuropathological variables in different brain regions in distinct groups of cases (all PSP stage > 2) defined based on HLA haplotypes (see manuscript). Color coding indicates the different brain regions. Abbreviations: GP, globus pallidus; CI, capsula interna; PUT, putamen; MCX, motor cortex; MWM, motor white matter; TDM, thalamus dorsomedial nucleus; STN, subthalamic nucleus; RN, red nucleus; MP, midbrain peduncles; SN, substantia nigra; OC, oculomotor complex.

| | | GROUP 1 | | | | | GROUP 2 | | | | | GROUP 3 | | | | | GROUP 4 | |
|---|---|---|---|---|---|---|---|---|---|---|---|---|---|---|---|---|---|---|
| | | N | Mean | SD | SE | | N | Mean | SD | SE | | N | Mean | SD | SE | | N | Mean |
| CD20/CD8 | PUT | 9 | 0.27 | 0.11 | 0.04 | PUT | 9 | 0.54 | 0.27 | 0.09 | SN | 7 | 1.51 | 3.47 | 1.31 | RN | 1 | 0.9749 |
| | MCX | 8 | 0.26 | 0.23 | 0.08 | GP | 6 | 0.27 | 0.12 | 0.05 | PUT | 6 | 0.23 | 0.13 | 0.05 | MCX | 1 | 0.1319 |
| | SN | 9 | 0.21 | 0.11 | 0.04 | CI | 8 | 0.22 | 0.33 | 0.12 | MP | 7 | 0.14 | 0.28 | 0.11 | SN | 1 | 0.0808 |
| | GP | 9 | 0.16 | 0.11 | 0.04 | SN | 9 | 0.21 | 0.13 | 0.04 | MCX | 5 | 0.13 | 0.13 | 0.06 | PUT | 1 | 0.0689 |
| | STN | 8 | 0.06 | 0.05 | 0.02 | MCX | 6 | 0.19 | 0.10 | 0.04 | GP | 6 | 0.11 | 0.05 | 0.02 | MWM | 1 | 0.0568 |
| | MWM | 8 | 0.06 | 0.03 | 0.01 | MWM | 6 | 0.09 | 0.10 | 0.04 | RN | 7 | 0.10 | 0.16 | 0.06 | TDM | 1 | 0.0501 |
| | TDM | 9 | 0.06 | 0.05 | 0.02 | RN | 9 | 0.09 | 0.14 | 0.05 | STN | 6 | 0.09 | 0.09 | 0.04 | MP | 1 | 0.0430 |
| | OC | 8 | 0.05 | 0.06 | 0.02 | OC | 8 | 0.09 | 0.11 | 0.04 | TDM | 7 | 0.09 | 0.08 | 0.03 | GP | 1 | 0.0211 |
| | RN | 9 | 0.04 | 0.05 | 0.02 | TDM | 9 | 0.08 | 0.05 | 0.02 | MWM | 5 | 0.05 | 0.06 | 0.03 | CI | 1 | 0.0136 |
| | MP | 9 | 0.03 | 0.02 | 0.01 | MP | 9 | 0.05 | 0.06 | 0.02 | OC | 7 | 0.04 | 0.08 | 0.03 | STN | 1 | 0.0000 |
| | CI | 9 | 0.03 | 0.02 | 0.01 | STN | 9 | 0.05 | 0.05 | 0.02 | CI | 5 | 0.04 | 0.04 | 0.02 | OC | 1 | 0.0000 |
| AT8/CD8 | MCX | 8 | 3.52 | 4.46 | 1.58 | MCX | 6 | 1.82 | 1.94 | 0.79 | MCX | 5 | 0.89 | 0.77 | 0.34 | MCX | 1 | 4.2124 |
| | OC | 8 | 0.87 | 1.06 | 0.38 | OC | 8 | 1.14 | 1.79 | 0.63 | CI | 5 | 0.37 | 0.45 | 0.20 | PUT | 1 | 0.6127 |
| | TDM | 9 | 0.53 | 0.40 | 0.13 | PUT | 9 | 0.77 | 0.71 | 0.24 | SN | 7 | 0.34 | 0.28 | 0.11 | RN | 1 | 0.5615 |
| | CI | 9 | 0.50 | 0.55 | 0.18 | RN | 9 | 0.73 | 0.53 | 0.18 | STN | 6 | 0.33 | 0.40 | 0.16 | TDM | 1 | 0.4921 |
| | SN | 9 | 0.46 | 0.51 | 0.17 | SN | 9 | 0.59 | 0.48 | 0.16 | RN | 7 | 0.33 | 0.54 | 0.20 | OC | 1 | 0.3460 |
| | STN | 8 | 0.43 | 0.57 | 0.20 | TDM | 9 | 0.57 | 0.48 | 0.16 | OC | 7 | 0.32 | 0.27 | 0.10 | CI | 1 | 0.3098 |
| | PUT | 9 | 0.43 | 0.39 | 0.13 | GP | 6 | 0.53 | 0.63 | 0.26 | TDM | 7 | 0.31 | 0.26 | 0.10 | MWM | 1 | 0.2456 |
| | GP | 9 | 0.34 | 0.36 | 0.12 | CI | 8 | 0.39 | 0.31 | 0.11 | PUT | 6 | 0.28 | 0.33 | 0.13 | GP | 1 | 0.2113 |
| | MWM | 8 | 0.33 | 0.33 | 0.12 | STN | 9 | 0.36 | 0.47 | 0.16 | GP | 6 | 0.23 | 0.26 | 0.11 | SN | 1 | 0.1999 |
| | MP | 9 | 0.27 | 0.27 | 0.09 | MP | 9 | 0.27 | 0.32 | 0.11 | MP | 7 | 0.17 | 0.21 | 0.08 | STN | 1 | 0.0932 |
| | RN | 9 | 0.23 | 0.23 | 0.08 | MWM | 6 | 0.15 | 0.13 | 0.05 | MWM | 5 | 0.15 | 0.14 | 0.06 | MP | 1 | 0.0711 |
| AT8/CD4 | OC | 8 | 2.56 | 2.88 | 1.02 | OC | 8 | 3.45 | 5.83 | 2.06 | MCX | 5 | 3.55 | 3.06 | 1.37 | MCX | 1 | 5.3213 |
| | MCX | 8 | 2.25 | 1.24 | 0.44 | TDM | 9 | 2.47 | 3.33 | 1.11 | TDM | 7 | 1.64 | 2.12 | 0.80 | OC | 1 | 2.8810 |
| | TDM | 9 | 1.40 | 1.85 | 0.62 | RN | 9 | 2.24 | 2.39 | 0.80 | OC | 7 | 1.32 | 1.56 | 0.59 | CI | 1 | 2.1665 |
| | CI | 9 | 1.01 | 1.01 | 0.34 | MP | 9 | 1.51 | 2.22 | 0.74 | CI | 5 | 0.87 | 0.49 | 0.22 | TDM | 1 | 1.6789 |
| | MP | 9 | 0.99 | 1.22 | 0.41 | CI | 8 | 1.46 | 2.34 | 0.83 | STN | 6 | 0.77 | 0.75 | 0.30 | STN | 1 | 0.8100 |
| | RN | 9 | 0.90 | 1.06 | 0.35 | MCX | 6 | 1.14 | 0.69 | 0.28 | GP | 6 | 0.72 | 0.64 | 0.26 | PUT | 1 | 0.7217 |
| | MWM | 8 | 0.70 | 0.82 | 0.29 | MWM | 6 | 1.02 | 1.95 | 0.80 | MWM | 5 | 0.59 | 0.45 | 0.20 | RN | 1 | 0.6752 |
| | GP | 9 | 0.52 | 0.45 | 0.15 | STN | 9 | 0.89 | 1.19 | 0.40 | RN | 7 | 0.57 | 0.40 | 0.15 | GP | 1 | 0.6274 |
| | STN | 8 | 0.50 | 0.59 | 0.21 | SN | 9 | 0.77 | 1.03 | 0.34 | MP | 7 | 0.51 | 0.44 | 0.17 | MWM | 1 | 0.3135 |
| | SN | 9 | 0.39 | 0.29 | 0.10 | PUT | 9 | 0.64 | 0.68 | 0.23 | PUT | 6 | 0.45 | 0.42 | 0.17 | MP | 1 | 0.1857 |
| | PUT | 9 | 0.34 | 0.55 | 0.18 | GP | 6 | 0.36 | 0.40 | 0.16 | SN | 7 | 0.19 | 0.17 | 0.07 | SN | 1 | 0.1718 |
| MG/CD8 | MCX | 8 | 2.92 | 2.69 | 0.95 | MCX | 6 | 1.89 | 2.00 | 0.82 | MP | 7 | 1.74 | 3.91 | 1.48 | MCX | 1 | 0.7870 |
| | CI | 9 | 1.84 | 1.03 | 0.34 | RN | 9 | 1.24 | 1.52 | 0.51 | SN | 7 | 1.03 | 1.74 | 0.66 | CI | 1 | 0.6903 |
| | PUT | 9 | 1.38 | 0.90 | 0.30 | TDM | 9 | 1.18 | 1.89 | 0.63 | STN | 6 | 0.76 | 1.05 | 0.43 | PUT | 1 | 0.6475 |
| | TDM | 9 | 1.23 | 0.80 | 0.27 | GP | 6 | 1.06 | 1.43 | 0.58 | RN | 7 | 0.74 | 1.40 | 0.53 | RN | 1 | 0.6249 |
| | MWM | 8 | 1.20 | 0.97 | 0.34 | OC | 8 | 0.99 | 0.95 | 0.33 | CI | 5 | 0.56 | 0.69 | 0.31 | MP | 1 | 0.2912 |
| | OC | 8 | 1.11 | 1.38 | 0.49 | PUT | 9 | 0.93 | 1.75 | 0.58 | TDM | 7 | 0.42 | 0.20 | 0.07 | SN | 1 | 0.2604 |
| | MP | 9 | 0.98 | 0.70 | 0.23 | MP | 9 | 0.91 | 0.99 | 0.33 | MCX | 5 | 0.41 | 0.22 | 0.10 | GP | 1 | 0.2424 |
| | GP | 9 | 0.89 | 0.96 | 0.32 | SN | 9 | 0.84 | 0.77 | 0.26 | GP | 6 | 0.30 | 0.37 | 0.15 | TDM | 1 | 0.2173 |
| | STN | 8 | 0.88 | 1.24 | 0.44 | CI | 8 | 0.79 | 0.69 | 0.24 | OC | 7 | 0.25 | 0.13 | 0.05 | MWM | 1 | 0.1758 |
| | SN | 9 | 0.79 | 0.78 | 0.26 | MWM | 6 | 0.63 | 0.38 | 0.16 | MWM | 5 | 0.24 | 0.15 | 0.07 | OC | 1 | 0.1183 |
| | RN | 9 | 0.46 | 0.41 | 0.14 | STN | 9 | 0.41 | 0.55 | 0.18 | PUT | 6 | 0.21 | 0.17 | 0.07 | STN | 1 | 0.0492 |
| MG/CD4 | CI | 9 | 3.50 | 2.28 | 0.76 | TDM | 9 | 3.27 | 7.02 | 2.34 | MP | 7 | 2.12 | 2.44 | 0.92 | CI | 1 | 4.8268 |
| | OC | 8 | 3.01 | 3.44 | 1.22 | MP | 9 | 3.04 | 3.51 | 1.17 | TDM | 7 | 1.65 | 1.63 | 0.61 | MCX | 1 | 0.9942 |
| | MP | 9 | 3.01 | 2.21 | 0.74 | MWM | 6 | 2.27 | 3.67 | 1.50 | MCX | 5 | 1.56 | 0.82 | 0.37 | OC | 1 | 0.9846 |
| | TDM | 9 | 2.45 | 1.65 | 0.55 | OC | 8 | 1.99 | 1.89 | 0.67 | STN | 6 | 1.37 | 1.38 | 0.56 | PUT | 1 | 0.7627 |
| | MWM | 8 | 2.32 | 1.50 | 0.53 | RN | 9 | 1.86 | 1.39 | 0.46 | CI | 5 | 1.30 | 0.50 | 0.23 | MP | 1 | 0.7601 |
| | MCX | 8 | 2.16 | 1.25 | 0.44 | CI | 8 | 1.71 | 2.07 | 0.73 | MWM | 5 | 0.99 | 0.76 | 0.34 | RN | 1 | 0.7515 |
| | RN | 9 | 1.35 | 0.88 | 0.29 | MCX | 6 | 1.12 | 0.77 | 0.31 | OC | 7 | 0.96 | 0.68 | 0.26 | TDM | 1 | 0.7412 |
| | GP | 9 | 1.26 | 1.03 | 0.34 | STN | 9 | 0.87 | 0.98 | 0.33 | RN | 7 | 0.89 | 0.47 | 0.18 | GP | 1 | 0.7198 |
| | STN | 8 | 1.00 | 1.16 | 0.41 | SN | 9 | 0.69 | 0.71 | 0.24 | GP | 6 | 0.86 | 0.76 | 0.31 | STN | 1 | 0.4281 |
| | PUT | 9 | 0.81 | 0.27 | 0.09 | GP | 6 | 0.59 | 0.44 | 0.18 | PUT | 6 | 0.33 | 0.18 | 0.07 | MWM | 1 | 0.2245 |
| | SN | 9 | 0.58 | 0.41 | 0.14 | PUT | 9 | 0.38 | 0.26 | 0.09 | SN | 7 | 0.31 | 0.23 | 0.09 | SN | 1 | 0.2238 |
| CD8/CD4 | OC | 8 | 4.09 | 4.25 | 1.50 | MP | 9 | 6.72 | 8.36 | 2.79 | MP | 7 | 7.35 | 8.30 | 3.14 | STN | 1 | 8.6945 |
| | RN | 9 | 4.07 | 3.36 | 1.12 | TDM | 9 | 6.45 | 10.75 | 3.58 | MWM | 5 | 5.76 | 6.16 | 2.75 | OC | 1 | 8.3260 |
| | MP | 9 | 4.02 | 3.28 | 1.09 | MWM | 6 | 5.65 | 9.74 | 3.98 | TDM | 7 | 5.14 | 5.12 | 1.94 | CI | 1 | 6.9925 |
| | MWM | 8 | 3.03 | 2.57 | 0.91 | RN | 9 | 4.46 | 4.22 | 1.41 | RN | 7 | 4.98 | 3.93 | 1.49 | TDM | 1 | 3.4118 |
| | CI | 9 | 2.90 | 2.69 | 0.90 | STN | 9 | 3.59 | 3.30 | 1.10 | OC | 7 | 4.24 | 3.11 | 1.17 | GP | 1 | 2.9689 |
| | TDM | 9 | 2.60 | 1.59 | 0.53 | CI | 8 | 3.25 | 4.14 | 1.46 | CI | 5 | 4.18 | 2.05 | 0.92 | MP | 1 | 2.6101 |
| | STN | 8 | 2.52 | 2.52 | 0.89 | OC | 8 | 2.98 | 2.48 | 0.88 | MCX | 5 | 4.13 | 1.82 | 0.81 | MWM | 1 | 1.2767 |
| | MCX | 8 | 2.30 | 2.27 | 0.80 | SN | 9 | 1.45 | 1.29 | 0.43 | GP | 6 | 3.38 | 1.23 | 0.50 | MCX | 1 | 1.2632 |
| | GP | 9 | 2.02 | 0.97 | 0.32 | MCX | 6 | 1.29 | 1.44 | 0.59 | STN | 6 | 3.31 | 2.14 | 0.87 | RN | 1 | 1.2026 |
| | SN | 9 | 1.25 | 1.22 | 0.41 | GP | 6 | 1.08 | 0.53 | 0.22 | PUT | 6 | 1.96 | 0.72 | 0.29 | PUT | 1 | 1.1779 |
| | PUT | 9 | 0.99 | 0.77 | 0.26 | PUT | 9 | 0.98 | 0.43 | 0.14 | SN | 7 | 1.04 | 1.34 | 0.51 | SN | 1 | 0.8595 |

**Table S5**. Mann-Whitney test results comparing neuropathological variables in different brain regions in distinct groups of cases defined based on HLA haplotypes (see manuscript). Red colored box and text indicates p value <0.05 and blue colored text indicates p< 0.01.

| Group 1 | | | | | | | | | | | |
|---|---|---|---|---|---|---|---|---|---|---|---|
| AT8 | | MICROGLIA | | CD3 | | CD4 | | CD8 | | CD20 | |
| Region | p | Region | p | Region | p | Region | p | Region | p | Region | p |
| PUT-TDM | 0.550 | MCx-PUT | 0.943 | TDM-MCx | 0.581 | MCx-TDM | 0.355 | MCx-TDM | 0.566 | CI-TDM | 0.919 |
| PUT-MP | 0.427 | MCx-TDM | 0.732 | TDM-CI | 0.190 | MCx-CI | 0.242 | MCx-PUT | 0.317 | CI-OC | 0.729 |
| PUT-MWM | 0.299 | MCx-OC | 0.020 | TDM-PUT | 0.099 | MCx-MP | 0.136 | MCx-CI | 0.112 | CI-RN | 0.308 |
| PUT-CI | 0.194 | MCx-MP | 0.018 | TDM-OC | 0.061 | MCx-MWM | 0.093 | MCx-OC | 0.093 | CI-STN | 0.274 |
| PUT-GP | 0.130 | MCx-MWM | 0.015 | TDM-MWM | 0.056 | MCx-OC | 0.071 | MCx-GP | 0.022 | CI-MP | 0.218 |
| PUT-MCx | 0.008 | MCx-GP | 0.008 | TDM-MP | 0.041 | MCx-PUT | 0.008 | MCx-MWM | 0.010 | CI-MCx | 0.143 |
| PUT-STN | 0.003 | MCx-SN | 0.002 | TDM-GP | 0.012 | MCx-GP | 0.005 | MCx-MP | 0.001 | CI-MWM | 0.043 |
| PUT-SN | 0.001 | MCx-CI | 0.001 | TDM-STN | 0.001 | MCx-STN | 0.003 | MCx-SN | 0.000 | CI-GP | 0.005 |
| PUT-RN | 0.001 | MCx-STN | 0.002 | TDM-SN | 0.000 | MCx-RN | 0.001 | MCx-STN | 0.000 | CI-PUT | 0.001 |
| PUT-OC | 0.000 | MCx-RN | 0.000 | TDM-RN | 0.000 | MCx-SN | 0.000 | MCx-RN | 0.000 | CI-SN | 0.000 |
| TDM-MP | 0.844 | PUT-TDM | 0.780 | MCx-CI | 0.471 | TDM-CI | 0.800 | TDM-PUT | 0.660 | TDM-OC | 0.806 |
| TDM-MWM | 0.647 | PUT-OC | 0.020 | MCx-PUT | 0.294 | TDM-MP | 0.559 | TDM-CI | 0.294 | TDM-RN | 0.358 |
| TDM-CI | 0.483 | PUT-MP | 0.018 | MCx-OC | 0.207 | TDM-MWM | 0.422 | TDM-OC | 0.253 | TDM-STN | 0.319 |
| TDM-GP | 0.360 | PUT-MWM | 0.015 | MCx-MWM | 0.187 | TDM-OC | 0.365 | TDM-GP | 0.077 | TDM-MP | 0.258 |
| TDM-MCx | 0.037 | PUT-GP | 0.008 | MCx-MP | 0.154 | TDM-PUT | 0.078 | TDM-MWM | 0.037 | TDM-MCx | 0.172 |
| TDM-STN | 0.018 | PUT-SN | 0.002 | MCx-GP | 0.058 | TDM-GP | 0.050 | TDM-MP | 0.007 | TDM-MWM | 0.054 |
| TDM-SN | 0.007 | PUT-CI | 0.001 | MCx-STN | 0.005 | TDM-STN | 0.030 | TDM-SN | 0.003 | TDM-GP | 0.007 |
| TDM-RN | 0.007 | PUT-STN | 0.001 | MCx-SN | 0.001 | TDM-RN | 0.017 | TDM-STN | 0.002 | TDM-PUT | 0.002 |
| TDM-OC | 0.000 | PUT-RN | 0.000 | MCx-RN | 0.000 | TDM-SN | 0.000 | TDM-RN | 0.000 | TDM-SN | 0.000 |
| MP-MWM | 0.789 | TDM-OC | 0.041 | CI-PUT | 0.735 | CI-MP | 0.741 | PUT-CI | 0.542 | OC-RN | 0.501 |
| MP-CI | 0.614 | TDM-MP | 0.037 | CI-OC | 0.577 | CI-MWM | 0.578 | PUT-OC | 0.483 | OC-STN | 0.449 |
| MP-GP | 0.473 | TDM-MWM | 0.030 | CI-MWM | 0.524 | CI-OC | 0.515 | PUT-GP | 0.184 | OC-MP | 0.376 |
| MP-MCx | 0.057 | TDM-GP | 0.018 | CI-MP | 0.467 | CI-PUT | 0.132 | PUT-MWM | 0.097 | OC-MCx | 0.260 |
| MP-STN | 0.029 | TDM-SN | 0.005 | CI-GP | 0.226 | CI-GP | 0.087 | PUT-MP | 0.023 | OC-MWM | 0.091 |
| MP-SN | 0.013 | TDM-CI | 0.003 | CI-STN | 0.031 | CI-STN | 0.054 | PUT-SN | 0.010 | OC-GP | 0.014 |
| MP-RN | 0.012 | TDM-STN | 0.004 | CI-SN | 0.005 | CI-RN | 0.033 | PUT-STN | 0.009 | OC-PUT | 0.004 |
| MP-OC | 0.001 | TDM-RN | 0.000 | CI-RN | 0.002 | CI-SN | 0.000 | PUT-RN | 0.000 | OC-SN | 0.001 |
| MWM-CI | 0.824 | OC-MP | 0.973 | PUT-OC | 0.826 | MP-MWM | 0.813 | CI-OC | 0.926 | RN-STN | 0.917 |
| MWM-GP | 0.668 | OC-MWM | 0.855 | PUT-MWM | 0.757 | MP-OC | 0.748 | CI-GP | 0.472 | RN-MP | 0.832 |
| MWM-MCx | 0.113 | OC-GP | 0.748 | PUT-MP | 0.697 | MP-PUT | 0.240 | CI-MWM | 0.285 | RN-MCx | 0.635 |
| MWM-STN | 0.061 | OC-SN | 0.451 | PUT-GP | 0.383 | MP-GP | 0.168 | CI-MP | 0.096 | RN-MWM | 0.299 |
| MWM-SN | 0.032 | OC-CI | 0.365 | PUT-STN | 0.067 | MP-STN | 0.109 | CI-SN | 0.051 | RN-GP | 0.073 |
| MWM-RN | 0.031 | OC-STN | 0.352 | PUT-SN | 0.014 | MP-RN | 0.072 | CI-STN | 0.043 | RN-PUT | 0.028 |
| MWM-OC | 0.002 | OC-RN | 0.104 | PUT-RN | 0.006 | MP-SN | 0.001 | CI-RN | 0.002 | RN-SN | 0.006 |
| CI-GP | 0.831 | MP-MWM | 0.881 | OC-MWM | 0.924 | MWM-OC | 0.939 | OC-GP | 0.531 | STN-MP | 0.920 |
| CI-MCx | 0.158 | MP-GP | 0.774 | OC-MP | 0.866 | MWM-PUT | 0.365 | OC-MWM | 0.328 | STN-MCx | 0.719 |
| CI-STN | 0.087 | MP-SN | 0.472 | OC-GP | 0.515 | MWM-GP | 0.270 | OC-MP | 0.116 | STN-MWM | 0.364 |
| CI-SN | 0.047 | MP-CI | 0.383 | OC-STN | 0.105 | MWM-STN | 0.184 | OC-SN | 0.063 | STN-GP | 0.102 |
| CI-RN | 0.045 | MP-STN | 0.369 | OC-SN | 0.025 | MWM-RN | 0.130 | OC-STN | 0.054 | STN-PUT | 0.043 |
| CI-OC | 0.003 | MP-RN | 0.112 | OC-RN | 0.011 | MWM-SN | 0.004 | OC-RN | 0.003 | STN-SN | 0.010 |
| GP-MCx | 0.229 | MWM-GP | 0.897 | MWM-MP | 0.945 | OC-PUT | 0.393 | GP-MWM | 0.711 | MP-MCx | 0.788 |
| GP-STN | 0.131 | MWM-SN | 0.584 | MWM-GP | 0.592 | OC-GP | 0.290 | GP-MP | 0.343 | MP-MWM | 0.405 |
| GP-SN | 0.077 | MWM-CI | 0.487 | MWM-STN | 0.139 | OC-STN | 0.197 | GP-SN | 0.217 | MP-GP | 0.113 |
| GP-RN | 0.074 | MWM-STN | 0.467 | MWM-SN | 0.038 | OC-RN | 0.139 | GP-STN | 0.186 | MP-PUT | 0.048 |
| GP-OC | 0.007 | MWM-RN | 0.163 | MWM-RN | 0.017 | OC-SN | 0.004 | GP-RN | 0.021 | MP-SN | 0.011 |
| MCx-STN | 0.733 | GP-SN | 0.666 | MP-GP | 0.630 | PUT-GP | 0.839 | MWM-MP | 0.583 | MCx-MWM | 0.584 |
| MCx-SN | 0.608 | GP-CI | 0.559 | MP-STN | 0.146 | PUT-STN | 0.644 | MWM-SN | 0.408 | MCx-GP | 0.205 |
| MCx-RN | 0.596 | GP-STN | 0.536 | MP-SN | 0.038 | PUT-RN | 0.531 | MWM-STN | 0.355 | MCx-PUT | 0.098 |
| MCx-OC | 0.154 | GP-RN | 0.193 | MP-RN | 0.017 | PUT-SN | 0.045 | MWM-RN | 0.061 | MCx-SN | 0.028 |
| STN-SN | 0.886 | SN-CI | 0.879 | GP-STN | 0.324 | GP-STN | 0.791 | MP-SN | 0.774 | MWM-GP | 0.482 |
| STN-RN | 0.873 | SN-STN | 0.841 | GP-SN | 0.112 | GP-RN | 0.672 | MP-STN | 0.688 | MWM-PUT | 0.276 |
| STN-OC | 0.306 | SN-RN | 0.383 | GP-RN | 0.057 | GP-SN | 0.072 | MP-RN | 0.173 | MWM-SN | 0.102 |
| SN-RN | 0.986 | CI-STN | 0.958 | STN-SN | 0.578 | STN-RN | 0.884 | SN-STN | 0.902 | GP-PUT | 0.691 |
| SN-OC | 0.347 | CI-RN | 0.472 | STN-RN | 0.390 | STN-SN | 0.138 | SN-RN | 0.283 | GP-SN | 0.337 |
| RN-OC | 0.356 | STN-RN | 0.519 | SN-RN | 0.754 | RN-SN | 0.168 | STN-RN | 0.358 | PUT-SN | 0.573 |

| Group 2 | | | | | | | | | | | |
|---|---|---|---|---|---|---|---|---|---|---|---|
| **AT8** | | **MICROGLIA** | | **CD3** | | **CD4** | | **CD8** | | **CD20** | |
| Region | p | Region | p | Region | p | Region | p | Region | p | Region | p |
| MP-MWM | 0.550 | MCx-PUT | 0.768 | MCx-TDM | 0.929 | MCx-MP | 0.939 | MCx-TDM | 0.174 | RN-TDM | 0.745 |
| MP-CI | 0.325 | MCx-TDM | 0.500 | MCx-MP | 0.859 | MCx-OC | 0.816 | MCx-OC | 0.087 | RN-MP | 0.725 |
| MP-TDM | 0.188 | MCx-OC | 0.349 | MCx-CI | 0.534 | MCx-TDM | 0.696 | MCx-PUT | 0.075 | RN-OC | 0.491 |
| MP-PUT | 0.146 | MCx-MP | 0.118 | MCx-MWM | 0.433 | MCx-MWM | 0.348 | MCx-MP | 0.010 | RN-MCx | 0.434 |
| MP-MCx | 0.152 | MCx-RN | 0.094 | MCx-OC | 0.294 | MCx-CI | 0.271 | MCx-CI | 0.009 | RN-MWM | 0.232 |
| MP-RN | 0.030 | MCx-MWM | 0.112 | MCx-RN | 0.119 | MCx-RN | 0.225 | MCx-MWM | 0.005 | RN-STN | 0.139 |
| MP-GP | 0.040 | MCx-CI | 0.073 | MCx-STN | 0.051 | MCx-STN | 0.092 | MCx-RN | 0.001 | RN-CI | 0.036 |
| MP-OC | 0.026 | MCx-STN | 0.019 | MCx-PUT | 0.009 | MCx-PUT | 0.077 | MCx-GP | 0.001 | RN-PUT | 0.000 |
| MP-SN | 0.002 | MCx-SN | 0.015 | MCx-GP | 0.003 | MCx-GP | 0.018 | MCx-SN | 0.000 | RN-SN | 0.000 |
| MP-STN | 0.001 | MCx-GP | 0.016 | MCx-SN | 0.000 | MCx-SN | 0.003 | MCx-STN | 0.000 | RN-GP | 0.000 |
| MWM-CI | 0.778 | PUT-TDM | 0.672 | TDM-MP | 0.921 | MP-OC | 0.860 | TDM-OC | 0.670 | TDM-MP | 0.978 |
| MWM-TDM | 0.561 | PUT-OC | 0.471 | TDM-CI | 0.552 | MP-TDM | 0.725 | TDM-PUT | 0.639 | TDM-OC | 0.709 |
| MWM-PUT | 0.483 | PUT-MP | 0.157 | TDM-MWM | 0.441 | MP-MWM | 0.341 | TDM-MP | 0.176 | TDM-MCx | 0.623 |
| MWM-MCx | 0.446 | PUT-RN | 0.123 | TDM-OC | 0.285 | MP-CI | 0.252 | TDM-CI | 0.167 | TDM-MWM | 0.366 |
| MWM-RN | 0.178 | PUT-MWM | 0.147 | TDM-RN | 0.101 | MP-RN | 0.203 | TDM-MWM | 0.091 | TDM-STN | 0.248 |
| MWM-GP | 0.194 | PUT-CI | 0.093 | TDM-STN | 0.037 | MP-STN | 0.073 | TDM-RN | 0.019 | TDM-CI | 0.077 |
| MWM-OC | 0.154 | PUT-STN | 0.022 | TDM-PUT | 0.005 | MP-PUT | 0.058 | TDM-GP | 0.019 | TDM-PUT | 0.000 |
| MWM-SN | 0.031 | PUT-SN | 0.016 | TDM-GP | 0.001 | MP-GP | 0.011 | TDM-SN | 0.002 | TDM-SN | 0.000 |
| MWM-STN | 0.016 | PUT-GP | 0.019 | TDM-SN | 0.000 | MP-SN | 0.001 | TDM-STN | 0.000 | TDM-GP | 0.000 |
| CI-TDM | 0.739 | TDM-OC | 0.757 | MP-CI | 0.620 | OC-TDM | 0.869 | OC-PUT | 0.976 | MP-OC | 0.729 |
| CI-PUT | 0.639 | TDM-MP | 0.321 | MP-MWM | 0.495 | OC-MWM | 0.441 | OC-MP | 0.375 | MP-MCx | 0.640 |
| CI-MCx | 0.580 | TDM-RN | 0.263 | MP-OC | 0.331 | OC-CI | 0.350 | OC-CI | 0.361 | MP-MWM | 0.379 |
| CI-RN | 0.234 | TDM-MWM | 0.285 | MP-RN | 0.123 | OC-RN | 0.290 | OC-MWM | 0.205 | MP-STN | 0.259 |
| CI-GP | 0.256 | TDM-CI | 0.210 | MP-STN | 0.047 | OC-STN | 0.117 | OC-RN | 0.066 | MP-CI | 0.082 |
| CI-OC | 0.201 | TDM-STN | 0.063 | MP-PUT | 0.007 | OC-PUT | 0.097 | OC-GP | 0.059 | MP-PUT | 0.000 |
| CI-SN | 0.036 | TDM-SN | 0.048 | MP-GP | 0.002 | OC-GP | 0.021 | OC-SN | 0.011 | MP-SN | 0.000 |
| CI-STN | 0.018 | TDM-GP | 0.051 | MP-SN | 0.000 | OC-SN | 0.003 | OC-STN | 0.002 | MP-GP | 0.000 |
| TDM-PUT | 0.892 | OC-MP | 0.514 | CI-MWM | 0.812 | TDM-MWM | 0.524 | PUT-MP | 0.377 | OC-MCx | 0.885 |
| TDM-MCx | 0.799 | OC-RN | 0.438 | CI-OC | 0.623 | TDM-CI | 0.427 | PUT-CI | 0.362 | OC-MWM | 0.585 |
| TDM-RN | 0.391 | OC-MWM | 0.444 | CI-RN | 0.295 | TDM-RN | 0.357 | PUT-MWM | 0.204 | OC-STN | 0.455 |
| TDM-GP | 0.410 | OC-CI | 0.364 | CI-STN | 0.137 | TDM-STN | 0.149 | PUT-RN | 0.062 | OC-CI | 0.179 |
| TDM-OC | 0.340 | OC-STN | 0.135 | CI-PUT | 0.027 | TDM-PUT | 0.123 | PUT-GP | 0.056 | OC-PUT | 0.002 |
| TDM-SN | 0.077 | OC-SN | 0.108 | CI-GP | 0.008 | TDM-GP | 0.027 | PUT-SN | 0.010 | OC-SN | 0.002 |
| TDM-STN | 0.041 | OC-GP | 0.107 | CI-SN | 0.001 | TDM-SN | 0.003 | PUT-STN | 0.001 | OC-GP | 0.001 |
| PUT-MCx | 0.894 | MP-RN | 0.899 | MWM-OC | 0.834 | MWM-CI | 0.942 | MP-CI | 0.978 | MCx-MWM | 0.707 |
| PUT-RN | 0.470 | MP-MWM | 0.856 | MWM-RN | 0.485 | MWM-RN | 0.853 | MP-MWM | 0.631 | MCx-STN | 0.589 |
| PUT-GP | 0.485 | MP-CI | 0.794 | MWM-STN | 0.274 | MWM-STN | 0.513 | MP-RN | 0.325 | MCx-CI | 0.276 |
| PUT-OC | 0.410 | MP-STN | 0.386 | MWM-PUT | 0.082 | MWM-PUT | 0.458 | MP-GP | 0.279 | MCx-PUT | 0.006 |
| PUT-SN | 0.102 | MP-SN | 0.325 | MWM-GP | 0.029 | MWM-GP | 0.163 | MP-SN | 0.088 | MCx-SN | 0.006 |
| PUT-STN | 0.057 | MP-GP | 0.306 | MWM-SN | 0.005 | MWM-SN | 0.046 | MP-STN | 0.021 | MCx-GP | 0.004 |
| MCx-RN | 0.608 | RN-MWM | 0.945 | OC-RN | 0.600 | CI-RN | 0.899 | CI-MWM | 0.648 | MWM-STN | 0.897 |
| MCx-GP | 0.613 | RN-CI | 0.892 | OC-STN | 0.341 | CI-STN | 0.516 | CI-RN | 0.339 | MWM-CI | 0.498 |
| MCx-OC | 0.541 | RN-STN | 0.459 | OC-PUT | 0.098 | CI-PUT | 0.454 | CI-GP | 0.290 | MWM-PUT | 0.020 |
| MCx-SN | 0.184 | RN-SN | 0.391 | OC-GP | 0.034 | CI-GP | 0.143 | CI-SN | 0.093 | MWM-SN | 0.019 |
| MCx-STN | 0.117 | RN-GP | 0.365 | OC-SN | 0.005 | CI-SN | 0.032 | CI-STN | 0.023 | MWM-GP | 0.014 |
| RN-GP | 0.982 | MWM-CI | 0.958 | RN-STN | 0.658 | RN-STN | 0.601 | MWM-RN | 0.690 | STN-CI | 0.540 |
| RN-OC | 0.902 | MWM-STN | 0.553 | RN-PUT | 0.244 | RN-PUT | 0.534 | MWM-GP | 0.599 | STN-PUT | 0.014 |
| RN-SN | 0.362 | MWM-SN | 0.485 | RN-GP | 0.094 | RN-GP | 0.178 | MWM-SN | 0.296 | STN-SN | 0.013 |
| RN-STN | 0.237 | MWM-GP | 0.450 | RN-SN | 0.019 | RN-SN | 0.043 | MWM-STN | 0.115 | STN-GP | 0.010 |
| GP-OC | 0.925 | CI-STN | 0.546 | STN-PUT | 0.470 | STN-PUT | 0.921 | RN-GP | 0.871 | CI-PUT | 0.066 |
| GP-SN | 0.407 | CI-SN | 0.470 | STN-GP | 0.206 | STN-GP | 0.391 | RN-SN | 0.470 | CI-SN | 0.062 |
| GP-STN | 0.279 | CI-GP | 0.436 | STN-SN | 0.056 | STN-SN | 0.134 | RN-STN | 0.188 | CI-GP | 0.044 |
| OC-SN | 0.447 | STN-SN | 0.907 | PUT-GP | 0.556 | PUT-GP | 0.445 | GP-SN | 0.608 | PUT-SN | 0.978 |
| OC-STN | 0.306 | STN-GP | 0.831 | PUT-SN | 0.234 | PUT-SN | 0.162 | GP-STN | 0.285 | PUT-GP | 0.772 |
| SN-STN | 0.787 | SN-GP | 0.917 | GP-SN | 0.599 | GP-SN | 0.587 | SN-STN | 0.552 | SN-GP | 0.792 |

| Group 3 | | | | | | | | | | | |
|---|---|---|---|---|---|---|---|---|---|---|---|
| AT8 | | MICROGLIA | | CD3 | | CD4 | | CD8 | | CD20 | |
| Region | p | Region | p | Region | p | Region | p | Region | p | Region | p |
| TDM-MP | 0.906 | PUT-TDM | 0.646 | TDM-MCx | 0.824 | MCx-TDM | 0.484 | TDM-MCx | 0.657 | TDM-CI | 0.861 |
| TDM-PUT | 0.783 | PUT-MCx | 0.583 | TDM-CI | 0.309 | MCx-CI | 0.290 | TDM-PUT | 0.431 | TDM-OC | 0.554 |
| TDM-CI | 0.363 | PUT-GP | 0.116 | TDM-PUT | 0.115 | MCx-MP | 0.118 | TDM-CI | 0.207 | TDM-STN | 0.513 |
| TDM-MWM | 0.376 | PUT-CI | 0.058 | TDM-MP | 0.112 | MCx-PUT | 0.029 | TDM-SN | 0.022 | TDM-MP | 0.400 |
| TDM-GP | 0.284 | PUT-MP | 0.015 | TDM-OC | 0.040 | MCx-GP | 0.021 | TDM-GP | 0.025 | TDM-RN | 0.328 |
| TDM-SN | 0.093 | PUT-MWM | 0.024 | TDM-GP | 0.044 | MCx-OC | 0.012 | TDM-MP | 0.018 | TDM-MCx | 0.366 |
| TDM-RN | 0.038 | PUT-OC | 0.013 | TDM-STN | 0.014 | MCx-STN | 0.012 | TDM-STN | 0.004 | TDM-MWM | 0.197 |
| TDM-MCx | 0.038 | PUT-STN | 0.014 | TDM-MWM | 0.006 | MCx-MWM | 0.010 | TDM-OC | 0.003 | TDM-GP | 0.008 |
| TDM-OC | 0.007 | PUT-RN | 0.003 | TDM-RN | 0.002 | MCx-RN | 0.004 | TDM-MWM | 0.003 | TDM-PUT | 0.004 |
| TDM-STN | 0.005 | PUT-SN | 0.002 | TDM-SN | 0.000 | MCx-SN | 0.000 | TDM-RN | 0.001 | TDM-SN | 0.000 |
| MP-PUT | 0.875 | TDM-MCx | 0.897 | MCx-CI | 0.472 | TDM-CI | 0.679 | MCx-PUT | 0.783 | CI-OC | 0.695 |
| MP-CI | 0.425 | TDM-GP | 0.258 | MCx-PUT | 0.224 | TDM-MP | 0.344 | MCx-CI | 0.465 | CI-STN | 0.645 |
| MP-MWM | 0.437 | TDM-CI | 0.146 | MCx-MP | 0.220 | TDM-PUT | 0.103 | MCx-SN | 0.098 | CI-MP | 0.527 |
| MP-GP | 0.338 | TDM-MP | 0.047 | MCx-OC | 0.100 | TDM-GP | 0.075 | MCx-GP | 0.102 | CI-RN | 0.444 |
| MP-SN | 0.118 | TDM-MWM | 0.066 | MCx-GP | 0.101 | TDM-OC | 0.047 | MCx-MP | 0.086 | CI-MCx | 0.476 |
| MP-RN | 0.050 | TDM-OC | 0.042 | MCx-STN | 0.042 | TDM-STN | 0.045 | MCx-STN | 0.028 | CI-MWM | 0.277 |
| MP-MCx | 0.049 | TDM-STN | 0.044 | MCx-MWM | 0.018 | TDM-MWM | 0.039 | MCx-OC | 0.021 | CI-GP | 0.017 |
| MP-OC | 0.010 | TDM-RN | 0.013 | MCx-RN | 0.010 | TDM-RN | 0.016 | MCx-MWM | 0.021 | CI-PUT | 0.009 |
| MP-STN | 0.006 | TDM-SN | 0.007 | MCx-SN | 0.000 | TDM-SN | 0.000 | MCx-RN | 0.011 | CI-SN | 0.001 |
| PUT-CI | 0.519 | MCx-GP | 0.361 | CI-PUT | 0.620 | CI-MP | 0.621 | PUT-CI | 0.614 | OC-STN | 0.931 |
| PUT-MWM | 0.527 | MCx-CI | 0.226 | CI-MP | 0.611 | CI-PUT | 0.250 | PUT-SN | 0.131 | OC-MP | 0.803 |
| PUT-GP | 0.419 | MCx-MP | 0.093 | CI-OC | 0.342 | CI-GP | 0.187 | PUT-GP | 0.136 | OC-RN | 0.698 |
| PUT-SN | 0.160 | MCx-MWM | 0.113 | CI-GP | 0.335 | CI-OC | 0.136 | PUT-MP | 0.115 | OC-MCx | 0.715 |
| PUT-RN | 0.072 | MCx-OC | 0.084 | CI-STN | 0.169 | CI-STN | 0.126 | PUT-STN | 0.036 | OC-MWM | 0.453 |
| PUT-MCx | 0.068 | MCx-STN | 0.084 | CI-MWM | 0.081 | CI-MWM | 0.105 | PUT-OC | 0.026 | OC-GP | 0.037 |
| PUT-OC | 0.016 | MCx-RN | 0.032 | CI-RN | 0.053 | CI-RN | 0.058 | PUT-MWM | 0.027 | OC-PUT | 0.022 |
| PUT-STN | 0.010 | MCx-SN | 0.021 | CI-SN | 0.003 | CI-SN | 0.000 | PUT-RN | 0.014 | OC-SN | 0.002 |
| CI-MWM | 0.985 | GP-CI | 0.755 | PUT-MP | 0.990 | MP-PUT | 0.495 | CI-SN | 0.344 | STN-MP | 0.878 |
| CI-GP | 0.876 | GP-MP | 0.439 | PUT-OC | 0.636 | MP-GP | 0.382 | CI-GP | 0.342 | STN-RN | 0.775 |
| CI-SN | 0.481 | GP-MWM | 0.459 | PUT-GP | 0.614 | MP-OC | 0.300 | CI-MP | 0.313 | STN-MCx | 0.785 |
| CI-RN | 0.279 | GP-OC | 0.410 | PUT-STN | 0.352 | MP-STN | 0.273 | CI-STN | 0.126 | STN-MWM | 0.518 |
| CI-MCx | 0.242 | GP-STN | 0.395 | PUT-MWM | 0.182 | MP-MWM | 0.228 | CI-OC | 0.104 | STN-GP | 0.054 |
| CI-OC | 0.094 | GP-RN | 0.206 | PUT-RN | 0.134 | MP-RN | 0.145 | CI-MWM | 0.095 | STN-PUT | 0.034 |
| CI-STN | 0.063 | GP-SN | 0.149 | PUT-SN | 0.011 | MP-SN | 0.001 | CI-RN | 0.062 | STN-SN | 0.003 |
| MWM-GP | 0.897 | CI-MP | 0.653 | MP-OC | 0.646 | PUT-GP | 0.827 | SN-GP | 0.968 | MP-RN | 0.890 |
| MWM-SN | 0.516 | CI-MWM | 0.657 | MP-GP | 0.623 | PUT-OC | 0.723 | SN-MP | 0.948 | MP-MCx | 0.891 |
| MWM-RN | 0.313 | CI-OC | 0.617 | MP-STN | 0.358 | PUT-STN | 0.660 | SN-STN | 0.520 | MP-MWM | 0.601 |
| MWM-MCx | 0.270 | CI-STN | 0.590 | MP-MWM | 0.186 | PUT-MWM | 0.560 | SN-OC | 0.478 | MP-GP | 0.064 |
| MWM-OC | 0.116 | CI-RN | 0.346 | MP-RN | 0.138 | PUT-RN | 0.438 | SN-MWM | 0.407 | MP-PUT | 0.040 |
| MWM-STN | 0.080 | CI-SN | 0.263 | MP-SN | 0.012 | PUT-SN | 0.011 | SN-RN | 0.338 | MP-SN | 0.004 |
| GP-SN | 0.587 | MP-MWM | 0.975 | OC-GP | 0.960 | GP-OC | 0.903 | GP-MP | 0.982 | RN-MCx | 0.991 |
| GP-RN | 0.357 | MP-OC | 0.958 | OC-STN | 0.633 | GP-STN | 0.832 | GP-STN | 0.561 | RN-MWM | 0.691 |
| GP-MCx | 0.307 | MP-STN | 0.913 | OC-MWM | 0.366 | GP-MWM | 0.717 | GP-OC | 0.521 | RN-GP | 0.086 |
| GP-OC | 0.130 | MP-RN | 0.609 | OC-RN | 0.306 | GP-RN | 0.599 | GP-MWM | 0.444 | RN-PUT | 0.056 |
| GP-STN | 0.089 | MP-SN | 0.486 | OC-SN | 0.039 | GP-SN | 0.026 | GP-RN | 0.378 | RN-SN | 0.006 |
| SN-RN | 0.694 | MWM-OC | 0.987 | GP-STN | 0.681 | OC-STN | 0.921 | MP-STN | 0.562 | MCx-MWM | 0.721 |
| SN-MCx | 0.588 | MWM-STN | 0.944 | GP-MWM | 0.408 | OC-MWM | 0.796 | MP-OC | 0.520 | MCx-GP | 0.117 |
| SN-OC | 0.312 | MWM-RN | 0.663 | GP-RN | 0.350 | OC-RN | 0.674 | MP-MWM | 0.442 | MCx-PUT | 0.083 |
| SN-STN | 0.221 | MWM-SN | 0.546 | GP-SN | 0.054 | OC-SN | 0.028 | MP-RN | 0.372 | MCx-SN | 0.012 |
| RN-MCx | 0.855 | OC-STN | 0.953 | STN-MWM | 0.663 | STN-MWM | 0.873 | STN-OC | 0.970 | MWM-GP | 0.233 |
| RN-OC | 0.537 | OC-RN | 0.646 | STN-RN | 0.612 | STN-RN | 0.760 | STN-MWM | 0.833 | MWM-PUT | 0.178 |
| RN-STN | 0.398 | OC-SN | 0.520 | STN-SN | 0.133 | STN-SN | 0.045 | STN-RN | 0.781 | MWM-SN | 0.034 |
| MCx-OC | 0.703 | STN-RN | 0.702 | MWM-RN | 0.975 | MWM-RN | 0.901 | OC-MWM | 0.855 | GP-PUT | 0.905 |
| MCx-STN | 0.548 | STN-SN | 0.576 | MWM-SN | 0.328 | MWM-SN | 0.081 | OC-RN | 0.803 | GP-SN | 0.354 |
| OC-STN | 0.801 | RN-SN | 0.854 | RN-SN | 0.300 | RN-SN | 0.076 | MWM-RN | 0.964 | PUT-SN | 0.400 |

**Table S6**. Mann-Whitney test results comparing ratios of neuropathological variables in different brain regions in distinct groups of cases defined based on HLA haplotypes (see manuscript). Red colored box and text indicates p value <0.05 and blue colored text indicates p< 0.01.

| Group 1 | | | | | | | | | | | |
|---|---|---|---|---|---|---|---|---|---|---|---|
| CD20/CD8 | | AT8/CD8 | | AT8/CD4 | | MG/CD8 | | MG/CD4 | | CD8/CD4 | |
| Region | p | Region | p | Region | p | Region | p | Region | p | Region | p |
| CI-RN | 0.861 | CI-RN | 0.861 | PUT-SN | 0.407 | RN-STN | 0.579 | SN-PUT | 0.578 | PUT-SN | 0.707 |
| CI-MP | 0.672 | CI-MP | 0.672 | PUT-STN | 0.360 | RN-GP | 0.385 | SN-STN | 0.583 | PUT-MCx | 0.200 |
| CI-OC | 0.491 | CI-OC | 0.491 | PUT-GP | 0.245 | RN-SN | 0.381 | SN-GP | 0.153 | PUT-STN | 0.099 |
| CI-TDM | 0.305 | CI-TDM | 0.305 | PUT-MWM | 0.134 | RN-OC | 0.209 | SN-RN | 0.107 | PUT-GP | 0.064 |
| CI-STN | 0.189 | CI-STN | 0.189 | PUT-RN | 0.100 | RN-MP | 0.110 | SN-OC | 0.010 | PUT-CI | 0.057 |
| CI-MWM | 0.140 | CI-MWM | 0.140 | PUT-MP | 0.054 | RN-MWM | 0.070 | SN-MCx | 0.003 | PUT-TDM | 0.031 |
| CI-GP | 0.002 | CI-GP | 0.002 | PUT-CI | 0.030 | RN-TDM | 0.032 | SN-MWM | 0.003 | PUT-MWM | 0.022 |
| CI-SN | 0.000 | CI-SN | 0.000 | PUT-TDM | 0.012 | RN-PUT | 0.017 | SN-TDM | 0.002 | PUT-OC | 0.020 |
| CI-MCx | 0.000 | CI-MCx | 0.000 | PUT-OC | 0.003 | RN-MCx | 0.010 | SN-MP | 0.000 | PUT-RN | 0.003 |
| CI-PUT | 0.000 | CI-PUT | 0.000 | PUT-MCx | 0.000 | RN-CI | 0.002 | SN-CI | 0.000 | PUT-MP | 0.002 |
| RN-MP | 0.804 | RN-MP | 0.804 | SN-STN | 0.911 | STN-GP | 0.774 | PUT-STN | 0.992 | SN-MCx | 0.359 |
| RN-OC | 0.604 | RN-OC | 0.604 | SN-GP | 0.739 | STN-SN | 0.768 | PUT-GP | 0.383 | SN-STN | 0.199 |
| RN-TDM | 0.395 | RN-TDM | 0.395 | SN-MWM | 0.487 | STN-OC | 0.496 | PUT-RN | 0.291 | SN-GP | 0.139 |
| RN-STN | 0.253 | RN-STN | 0.253 | SN-RN | 0.414 | STN-MP | 0.319 | PUT-OC | 0.042 | SN-CI | 0.126 |
| RN-MWM | 0.191 | RN-MWM | 0.191 | SN-MP | 0.272 | STN-MWM | 0.223 | PUT-MCx | 0.017 | SN-TDM | 0.074 |
| RN-GP | 0.003 | RN-GP | 0.003 | SN-CI | 0.179 | STN-TDM | 0.128 | PUT-MWM | 0.015 | SN-MWM | 0.054 |
| RN-SN | 0.001 | RN-SN | 0.001 | SN-TDM | 0.092 | STN-PUT | 0.079 | PUT-TDM | 0.010 | SN-OC | 0.050 |
| RN-MCx | 0.000 | RN-MCx | 0.000 | SN-OC | 0.030 | STN-MCx | 0.050 | PUT-MP | 0.003 | SN-RN | 0.010 |
| RN-PUT | 0.000 | RN-PUT | 0.000 | SN-MCx | 0.001 | STN-CI | 0.014 | PUT-CI | 0.001 | SN-MP | 0.007 |
| MP-OC | 0.781 | MP-OC | 0.781 | STN-GP | 0.832 | GP-SN | 0.993 | STN-GP | 0.403 | MCx-STN | 0.720 |
| MP-TDM | 0.547 | MP-TDM | 0.547 | STN-MWM | 0.571 | GP-OC | 0.679 | STN-RN | 0.310 | MCx-GP | 0.604 |
| MP-STN | 0.367 | MP-STN | 0.367 | STN-RN | 0.496 | GP-MP | 0.465 | STN-OC | 0.049 | MCx-CI | 0.570 |
| MP-MWM | 0.286 | MP-MWM | 0.286 | STN-MP | 0.340 | GP-MWM | 0.333 | STN-MCx | 0.021 | MCx-TDM | 0.414 |
| MP-GP | 0.007 | MP-GP | 0.007 | STN-CI | 0.234 | GP-TDM | 0.203 | STN-MWM | 0.019 | MCx-MWM | 0.327 |
| MP-SN | 0.002 | MP-SN | 0.002 | STN-TDM | 0.128 | GP-PUT | 0.130 | STN-TDM | 0.013 | MCx-OC | 0.312 |
| MP-MCx | 0.001 | MP-MCx | 0.001 | STN-OC | 0.045 | GP-MCx | 0.083 | STN-MP | 0.004 | MCx-RN | 0.113 |
| MP-PUT | 0.000 | MP-PUT | 0.000 | STN-MCx | 0.002 | GP-CI | 0.025 | STN-CI | 0.001 | MCx-MP | 0.092 |
| OC-TDM | 0.758 | OC-TDM | 0.758 | GP-MWM | 0.710 | SN-OC | 0.686 | GP-RN | 0.854 | STN-GP | 0.881 |
| OC-STN | 0.543 | OC-STN | 0.543 | GP-RN | 0.629 | SN-MP | 0.470 | GP-OC | 0.235 | STN-CI | 0.842 |
| OC-MWM | 0.443 | OC-MWM | 0.443 | GP-MP | 0.444 | SN-MWM | 0.337 | GP-MCx | 0.124 | STN-TDM | 0.654 |
| OC-GP | 0.019 | OC-GP | 0.019 | GP-CI | 0.313 | SN-TDM | 0.206 | GP-MWM | 0.113 | STN-MWM | 0.534 |
| OC-SN | 0.005 | OC-SN | 0.005 | GP-TDM | 0.177 | SN-PUT | 0.132 | GP-TDM | 0.090 | STN-OC | 0.514 |
| OC-MCx | 0.004 | OC-MCx | 0.004 | GP-OC | 0.064 | SN-MCx | 0.084 | GP-MP | 0.034 | STN-RN | 0.224 |
| OC-PUT | 0.000 | OC-PUT | 0.000 | GP-MCx | 0.003 | SN-CI | 0.026 | GP-CI | 0.012 | STN-MP | 0.188 |
| TDM-STN | 0.751 | TDM-STN | 0.751 | MWM-RN | 0.922 | OC-MP | 0.767 | RN-OC | 0.312 | GP-CI | 0.959 |
| TDM-MWM | 0.630 | TDM-MWM | 0.630 | MWM-MP | 0.710 | OC-MWM | 0.589 | RN-MCx | 0.174 | GP-TDM | 0.758 |
| TDM-GP | 0.035 | TDM-GP | 0.035 | MWM-CI | 0.543 | OC-TDM | 0.411 | RN-MWM | 0.159 | GP-MWM | 0.625 |
| TDM-SN | 0.010 | TDM-SN | 0.010 | MWM-TDM | 0.348 | OC-PUT | 0.291 | RN-TDM | 0.131 | GP-OC | 0.602 |
| TDM-MCx | 0.008 | TDM-MCx | 0.008 | MWM-OC | 0.151 | OC-MCx | 0.199 | RN-MP | 0.052 | GP-RN | 0.272 |
| TDM-PUT | 0.001 | TDM-PUT | 0.001 | MWM-MCx | 0.011 | OC-CI | 0.078 | RN-CI | 0.020 | GP-MP | 0.230 |
| STN-MWM | 0.874 | STN-MWM | 0.874 | RN-MP | 0.778 | MP-MWM | 0.795 | OC-MCx | 0.734 | CI-TDM | 0.798 |
| STN-GP | 0.085 | STN-GP | 0.085 | RN-CI | 0.599 | MP-TDM | 0.587 | OC-MWM | 0.700 | CI-MWM | 0.660 |
| STN-SN | 0.030 | STN-SN | 0.030 | RN-TDM | 0.385 | MP-PUT | 0.434 | OC-TDM | 0.650 | CI-OC | 0.637 |
| STN-MCx | 0.023 | STN-MCx | 0.023 | RN-OC | 0.167 | MP-MCx | 0.306 | OC-MP | 0.383 | CI-RN | 0.295 |
| STN-PUT | 0.003 | STN-PUT | 0.003 | RN-MCx | 0.012 | MP-CI | 0.131 | OC-CI | 0.211 | CI-MP | 0.250 |
| MWM-GP | 0.119 | MWM-GP | 0.119 | MP-CI | 0.807 | MWM-TDM | 0.789 | MCx-MWM | 0.964 | TDM-MWM | 0.849 |
| MWM-SN | 0.045 | MWM-SN | 0.045 | MP-TDM | 0.558 | MWM-PUT | 0.617 | MCx-TDM | 0.917 | TDM-OC | 0.823 |
| MWM-MCx | 0.034 | MWM-MCx | 0.034 | MP-OC | 0.268 | MWM-MCx | 0.457 | MCx-MP | 0.601 | TDM-RN | 0.429 |
| MWM-PUT | 0.005 | MWM-PUT | 0.005 | MP-MCx | 0.026 | MWM-CI | 0.228 | MCx-CI | 0.368 | TDM-MP | 0.372 |
| GP-SN | 0.644 | GP-SN | 0.644 | CI-TDM | 0.732 | TDM-PUT | 0.811 | MWM-TDM | 0.955 | MWM-OC | 0.975 |
| GP-MCx | 0.532 | GP-MCx | 0.532 | CI-OC | 0.384 | TDM-MCx | 0.619 | MWM-MP | 0.634 | MWM-RN | 0.564 |
| GP-PUT | 0.194 | GP-PUT | 0.194 | CI-MCx | 0.046 | TDM-CI | 0.334 | MWM-CI | 0.393 | MWM-MP | 0.499 |
| SN-MCx | 0.860 | SN-MCx | 0.860 | TDM-OC | 0.589 | PUT-MCx | 0.791 | TDM-MP | 0.666 | OC-RN | 0.587 |
| SN-PUT | 0.402 | SN-PUT | 0.402 | TDM-MCx | 0.096 | PUT-CI | 0.467 | TDM-CI | 0.412 | OC-MP | 0.520 |
| MCx-PUT | 0.524 | MCx-PUT | 0.524 | OC-MCx | 0.274 | MCx-CI | 0.660 | MP-CI | 0.697 | RN-MP | 0.918 |

| Group 2 | | | | | | | | | | | |
|---|---|---|---|---|---|---|---|---|---|---|---|
| **CD20/CD8** | | **AT8/CD8** | | **AT8/CD4** | | **MG/CD8** | | **MG/CD4** | | **CD8/CD4** | |
| Region | p | Region | p | Region | p | Region | p | Region | p | Region | p |
| MP-STN | 0.930 | MWM-MP | 0.473 | MWM-MP | 0.473 | STN-PUT | 0.469 | PUT-GP | 0.456 | PUT-MCx | 0.964 |
| MP-RN | 0.638 | MWM-STN | 0.285 | MWM-STN | 0.285 | STN-MP | 0.208 | PUT-STN | 0.344 | PUT-GP | 0.793 |
| MP-OC | 0.490 | MWM-GP | 0.212 | MWM-GP | 0.212 | STN-GP | 0.207 | PUT-SN | 0.344 | PUT-SN | 0.688 |
| MP-MWM | 0.416 | MWM-CI | 0.169 | MWM-CI | 0.169 | STN-TDM | 0.154 | PUT-TDM | 0.064 | PUT-CI | 0.161 |
| MP-TDM | 0.230 | MWM-SN | 0.040 | MWM-SN | 0.040 | STN-SN | 0.153 | PUT-MCx | 0.059 | PUT-MWM | 0.142 |
| MP-CI | 0.094 | MWM-TDM | 0.031 | MWM-TDM | 0.031 | STN-MWM | 0.180 | PUT-OC | 0.023 | PUT-TDM | 0.090 |
| MP-MCx | 0.013 | MWM-PUT | 0.023 | MWM-PUT | 0.023 | STN-CI | 0.129 | PUT-CI | 0.018 | PUT-OC | 0.065 |
| MP-SN | 0.003 | MWM-OC | 0.019 | MWM-OC | 0.019 | STN-OC | 0.106 | PUT-MWM | 0.023 | PUT-STN | 0.056 |
| MP-GP | 0.001 | MWM-RN | 0.008 | MWM-RN | 0.008 | STN-RN | 0.074 | PUT-RN | 0.002 | PUT-RN | 0.029 |
| MP-PUT | 0.000 | MWM-MCx | 0.001 | MWM-MCx | 0.001 | STN-MCx | 0.007 | PUT-MP | 0.001 | PUT-MP | 0.014 |
| STN-RN | 0.702 | MP-STN | 0.695 | MP-STN | 0.695 | PUT-MP | 0.593 | GP-STN | 0.919 | MCx-GP | 0.843 |
| STN-OC | 0.545 | MP-GP | 0.516 | MP-GP | 0.516 | PUT-GP | 0.540 | GP-SN | 0.919 | MCx-SN | 0.754 |
| STN-MWM | 0.463 | MP-CI | 0.453 | MP-CI | 0.453 | PUT-TDM | 0.483 | GP-TDM | 0.361 | MCx-CI | 0.224 |
| STN-TDM | 0.266 | MP-SN | 0.134 | MP-SN | 0.134 | PUT-SN | 0.480 | GP-MCx | 0.296 | MCx-MWM | 0.194 |
| STN-CI | 0.112 | MP-TDM | 0.106 | MP-TDM | 0.106 | PUT-MWM | 0.488 | GP-OC | 0.189 | MCx-TDM | 0.141 |
| STN-MCx | 0.016 | MP-PUT | 0.083 | MP-PUT | 0.083 | PUT-CI | 0.414 | GP-CI | 0.159 | MCx-OC | 0.106 |
| STN-SN | 0.004 | MP-OC | 0.068 | MP-OC | 0.068 | PUT-OC | 0.361 | GP-MWM | 0.163 | MCx-STN | 0.096 |
| STN-GP | 0.002 | MP-RN | 0.030 | MP-RN | 0.030 | PUT-RN | 0.289 | GP-RN | 0.048 | MCx-RN | 0.056 |
| STN-PUT | 0.000 | MP-MCx | 0.006 | MP-MCx | 0.006 | PUT-MCx | 0.041 | GP-MP | 0.022 | MCx-MP | 0.032 |
| RN-OC | 0.815 | STN-GP | 0.765 | STN-GP | 0.765 | MP-GP | 0.893 | STN-OC | 0.177 | GP-SN | 0.923 |
| RN-MWM | 0.695 | STN-CI | 0.712 | STN-CI | 0.712 | MP-TDM | 0.868 | STN-RN | 0.036 | GP-CI | 0.315 |
| RN-TDM | 0.466 | STN-SN | 0.268 | STN-SN | 0.268 | MP-SN | 0.864 | SN-CI | 0.145 | GP-MWM | 0.271 |
| RN-CI | 0.224 | STN-TDM | 0.222 | STN-TDM | 0.222 | MP-MWM | 0.830 | SN-MP | 0.015 | GP-TDM | 0.209 |
| RN-MCx | 0.039 | STN-PUT | 0.179 | STN-PUT | 0.179 | MP-CI | 0.766 | STN-TDM | 0.363 | GP-OC | 0.159 |
| RN-SN | 0.013 | STN-OC | 0.149 | STN-OC | 0.149 | MP-OC | 0.694 | STN-CI | 0.145 | GP-STN | 0.147 |
| RN-GP | 0.006 | STN-RN | 0.076 | STN-RN | 0.076 | MP-RN | 0.599 | SN-OC | 0.177 | GP-RN | 0.090 |
| RN-PUT | 0.000 | STN-MCx | 0.016 | STN-MCx | 0.016 | MP-MCx | 0.118 | SN-RN | 0.036 | GP-MP | 0.054 |
| OC-MWM | 0.863 | GP-CI | 0.967 | GP-CI | 0.967 | GP-TDM | 0.988 | STN-SN | 1.000 | SN-CI | 0.312 |
| OC-TDM | 0.636 | GP-SN | 0.489 | GP-SN | 0.489 | GP-SN | 0.985 | SN-MWM | 0.153 | SN-MWM | 0.267 |
| OC-CI | 0.340 | GP-TDM | 0.427 | GP-TDM | 0.427 | GP-MWM | 0.941 | STN-MCx | 0.297 | SN-TDM | 0.195 |
| OC-MCx | 0.071 | GP-PUT | 0.367 | GP-PUT | 0.367 | GP-CI | 0.891 | STN-MP | 0.015 | SN-OC | 0.145 |
| OC-SN | 0.029 | GP-OC | 0.315 | GP-OC | 0.315 | GP-OC | 0.823 | SN-TDM | 0.363 | SN-STN | 0.130 |
| OC-GP | 0.013 | GP-RN | 0.197 | GP-RN | 0.197 | GP-RN | 0.737 | SN-MCx | 0.297 | SN-RN | 0.074 |
| OC-PUT | 0.000 | GP-MCx | 0.054 | GP-MCx | 0.054 | GP-MCx | 0.192 | STN-MWM | 0.153 | SN-MP | 0.041 |
| MWM-TDM | 0.795 | CI-SN | 0.481 | CI-SN | 0.481 | TDM-SN | 0.996 | TDM-MCx | 0.817 | CI-MWM | 0.862 |
| MWM-CI | 0.477 | CI-TDM | 0.414 | CI-TDM | 0.414 | TDM-MWM | 0.947 | TDM-OC | 0.639 | CI-TDM | 0.805 |
| MWM-MCx | 0.127 | CI-PUT | 0.351 | CI-PUT | 0.351 | TDM-CI | 0.891 | TDM-CI | 0.566 | CI-OC | 0.663 |
| MWM-SN | 0.067 | CI-OC | 0.297 | CI-OC | 0.297 | TDM-OC | 0.816 | TDM-MWM | 0.539 | CI-STN | 0.648 |
| MWM-GP | 0.030 | CI-RN | 0.176 | CI-RN | 0.176 | TDM-RN | 0.719 | TDM-RN | 0.234 | CI-RN | 0.471 |
| MWM-PUT | 0.001 | CI-MCx | 0.044 | CI-MCx | 0.044 | TDM-MCx | 0.157 | TDM-MP | 0.126 | CI-MP | 0.331 |
| TDM-CI | 0.611 | SN-TDM | 0.908 | SN-TDM | 0.908 | SN-MWM | 0.951 | MCx-OC | 0.844 | MWM-TDM | 0.961 |
| TDM-MCx | 0.158 | SN-PUT | 0.814 | SN-PUT | 0.814 | SN-CI | 0.895 | MCx-CI | 0.771 | MWM-OC | 0.818 |
| TDM-SN | 0.079 | SN-OC | 0.713 | SN-OC | 0.713 | SN-OC | 0.819 | MCx-MWM | 0.726 | MWM-STN | 0.808 |
| TDM-GP | 0.035 | SN-RN | 0.504 | SN-RN | 0.504 | SN-RN | 0.722 | MCx-RN | 0.405 | MWM-RN | 0.626 |
| TDM-PUT | 0.001 | SN-MCx | 0.156 | SN-MCx | 0.156 | SN-MCx | 0.158 | MCx-MP | 0.255 | MWM-MP | 0.473 |
| CI-MCx | 0.358 | TDM-PUT | 0.905 | TDM-PUT | 0.905 | MWM-CI | 0.953 | OC-CI | 0.918 | TDM-OC | 0.840 |
| CI-SN | 0.231 | TDM-OC | 0.798 | TDM-OC | 0.798 | MWM-OC | 0.885 | OC-MWM | 0.859 | TDM-STN | 0.828 |
| CI-GP | 0.109 | TDM-RN | 0.580 | TDM-RN | 0.580 | MWM-RN | 0.798 | OC-RN | 0.493 | TDM-RN | 0.625 |
| CI-PUT | 0.007 | TDM-MCx | 0.188 | TDM-MCx | 0.188 | MWM-MCx | 0.218 | OC-MP | 0.309 | TDM-MP | 0.455 |
| MCx-SN | 0.872 | PUT-OC | 0.889 | PUT-OC | 0.889 | CI-OC | 0.926 | CI-MWM | 0.934 | OC-STN | 0.993 |
| MCx-GP | 0.523 | PUT-RN | 0.665 | PUT-RN | 0.665 | CI-RN | 0.832 | CI-RN | 0.562 | OC-RN | 0.785 |
| MCx-PUT | 0.127 | PUT-MCx | 0.227 | PUT-MCx | 0.227 | CI-MCx | 0.208 | CI-MP | 0.362 | OC-MP | 0.601 |
| SN-GP | 0.590 | OC-RN | 0.779 | OC-RN | 0.779 | OC-RN | 0.907 | MWM-RN | 0.653 | STN-RN | 0.785 |
| SN-PUT | 0.127 | OC-MCx | 0.292 | OC-MCx | 0.292 | OC-MCx | 0.241 | MWM-MP | 0.450 | STN-MP | 0.596 |
| GP-PUT | 0.408 | RN-MCx | 0.412 | RN-MCx | 0.412 | RN-MCx | 0.274 | RN-MP | 0.733 | RN-MP | 0.796 |

| Group 3 | | | | | | | | | | | |
|---|---|---|---|---|---|---|---|---|---|---|---|
| **CD20/CD8** | | **AT8/CD8** | | **AT8/CD4** | | **MG/CD8** | | **MG/CD4** | | **CD8/CD4** | |
| Region | p | Region | p | Region | p | Region | p | Region | p | Region | p |
| OC-CI | 0.814 | MP-MWM | 0.755 | SN-PUT | 0.289 | PUT-GP | 0.832 | SN-PUT | 0.840 | SN-PUT | 0.382 |
| OC-MWM | 0.627 | MP-RN | 0.534 | SN-MP | 0.183 | PUT-MWM | 0.668 | SN-GP | 0.096 | SN-STN | 0.053 |
| OC-RN | 0.575 | MP-GP | 0.527 | SN-RN | 0.090 | PUT-OC | 0.530 | SN-OC | 0.035 | SN-GP | 0.036 |
| OC-MP | 0.314 | MP-PUT | 0.361 | SN-MWM | 0.116 | PUT-RN | 0.445 | SN-RN | 0.027 | SN-TDM | 0.017 |
| OC-STN | 0.265 | MP-STN | 0.248 | SN-GP | 0.075 | PUT-STN | 0.194 | SN-MWM | 0.039 | SN-OC | 0.015 |
| OC-TDM | 0.123 | MP-CI | 0.198 | SN-STN | 0.071 | PUT-CI | 0.192 | SN-STN | 0.016 | SN-MWM | 0.020 |
| OC-MCx | 0.078 | MP-TDM | 0.101 | SN-TDM | 0.024 | PUT-MP | 0.129 | SN-TDM | 0.007 | SN-MP | 0.010 |
| OC-GP | 0.052 | MP-OC | 0.090 | SN-OC | 0.013 | PUT-MCx | 0.080 | SN-MP | 0.002 | SN-RN | 0.009 |
| OC-SN | 0.002 | MP-SN | 0.076 | SN-CI | 0.017 | PUT-TDM | 0.032 | SN-CI | 0.003 | SN-MCx | 0.015 |
| OC-PUT | 0.002 | MP-MCx | 0.003 | SN-MCx | 0.000 | PUT-SN | 0.026 | SN-MCx | 0.001 | SN-CI | 0.014 |
| CI-MWM | 0.817 | MWM-RN | 0.798 | PUT-MP | 0.827 | GP-MWM | 0.821 | PUT-GP | 0.159 | PUT-STN | 0.307 |
| CI-RN | 0.782 | MWM-GP | 0.780 | PUT-RN | 0.569 | GP-OC | 0.683 | PUT-OC | 0.068 | PUT-GP | 0.240 |
| CI-MP | 0.494 | MWM-PUT | 0.591 | PUT-MWM | 0.585 | GP-RN | 0.586 | PUT-RN | 0.055 | PUT-TDM | 0.158 |
| CI-STN | 0.426 | MWM-STN | 0.447 | PUT-GP | 0.488 | GP-STN | 0.277 | PUT-MWM | 0.070 | PUT-OC | 0.143 |
| CI-TDM | 0.242 | MWM-CI | 0.366 | PUT-STN | 0.474 | GP-CI | 0.270 | PUT-STN | 0.034 | PUT-MWM | 0.149 |
| CI-MCx | 0.157 | MWM-TDM | 0.235 | PUT-TDM | 0.268 | GP-MP | 0.194 | PUT-TDM | 0.017 | PUT-MP | 0.110 |
| CI-GP | 0.120 | MWM-OC | 0.216 | PUT-OC | 0.188 | GP-MCx | 0.122 | PUT-MP | 0.005 | PUT-RN | 0.101 |
| CI-SN | 0.008 | MWM-SN | 0.190 | PUT-CI | 0.181 | GP-TDM | 0.054 | PUT-CI | 0.007 | PUT-MCx | 0.123 |
| CI-PUT | 0.008 | MWM-MCx | 0.016 | PUT-MCx | 0.008 | GP-SN | 0.045 | PUT-MCx | 0.003 | PUT-CI | 0.113 |
| MWM-RN | 0.979 | RN-GP | 0.972 | MP-RN | 0.715 | MWM-OC | 0.878 | GP-OC | 0.717 | STN-GP | 0.878 |
| MWM-MP | 0.665 | RN-PUT | 0.752 | MP-MWM | 0.721 | MWM-RN | 0.778 | GP-RN | 0.646 | STN-TDM | 0.726 |
| MWM-STN | 0.580 | RN-STN | 0.577 | MP-GP | 0.616 | MWM-STN | 0.418 | GP-MWM | 0.639 | STN-OC | 0.687 |
| MWM-TDM | 0.357 | RN-CI | 0.471 | MP-STN | 0.600 | MWM-CI | 0.401 | GP-STN | 0.479 | STN-MWM | 0.640 |
| MWM-MCx | 0.237 | RN-TDM | 0.308 | MP-TDM | 0.355 | MWM-MP | 0.318 | GP-TDM | 0.350 | STN-MP | 0.590 |
| MWM-GP | 0.189 | RN-OC | 0.283 | MP-OC | 0.253 | MWM-MCx | 0.206 | GP-MP | 0.179 | STN-RN | 0.563 |
| MWM-SN | 0.017 | RN-SN | 0.248 | MP-CI | 0.239 | MWM-TDM | 0.110 | GP-CI | 0.178 | STN-MCx | 0.570 |
| MWM-PUT | 0.016 | RN-MCx | 0.019 | MP-MCx | 0.012 | MWM-SN | 0.095 | GP-MCx | 0.112 | STN-CI | 0.542 |
| RN-MP | 0.656 | GP-PUT | 0.787 | RN-MWM | 0.981 | OC-RN | 0.887 | OC-RN | 0.919 | GP-TDM | 0.848 |
| RN-STN | 0.565 | GP-STN | 0.614 | RN-GP | 0.880 | OC-STN | 0.471 | OC-MWM | 0.888 | GP-OC | 0.808 |
| RN-TDM | 0.327 | GP-CI | 0.507 | RN-STN | 0.863 | OC-CI | 0.451 | OC-STN | 0.710 | GP-MWM | 0.748 |
| RN-MCx | 0.210 | GP-TDM | 0.345 | RN-TDM | 0.575 | OC-MP | 0.355 | OC-TDM | 0.552 | GP-MP | 0.704 |
| RN-GP | 0.161 | GP-OC | 0.319 | RN-OC | 0.437 | OC-MCx | 0.226 | OC-MP | 0.308 | GP-RN | 0.675 |
| RN-SN | 0.010 | GP-SN | 0.283 | RN-CI | 0.399 | OC-TDM | 0.114 | OC-CI | 0.295 | GP-MCx | 0.673 |
| RN-PUT | 0.010 | GP-MCx | 0.025 | RN-MCx | 0.029 | OC-SN | 0.096 | OC-MCx | 0.194 | GP-CI | 0.643 |
| MP-STN | 0.883 | PUT-STN | 0.815 | MWM-GP | 0.908 | RN-STN | 0.559 | RN-MWM | 0.962 | TDM-OC | 0.957 |
| MP-TDM | 0.593 | PUT-CI | 0.684 | MWM-STN | 0.891 | RN-CI | 0.532 | RN-STN | 0.783 | TDM-MWM | 0.880 |
| MP-MCx | 0.398 | PUT-TDM | 0.506 | MWM-TDM | 0.625 | RN-MP | 0.433 | RN-TDM | 0.622 | TDM-MP | 0.845 |
| MP-GP | 0.331 | PUT-OC | 0.474 | MWM-OC | 0.493 | RN-MCx | 0.279 | RN-MP | 0.358 | TDM-RN | 0.813 |
| MP-SN | 0.032 | PUT-SN | 0.427 | MWM-CI | 0.447 | RN-TDM | 0.150 | RN-CI | 0.340 | TDM-MCx | 0.799 |
| MP-PUT | 0.031 | PUT-MCx | 0.047 | MWM-MCx | 0.046 | RN-SN | 0.128 | RN-MCx | 0.228 | TDM-CI | 0.766 |
| STN-TDM | 0.714 | STN-CI | 0.854 | GP-STN | 0.983 | STN-CI | 0.947 | MWM-STN | 0.837 | OC-MWM | 0.919 |
| STN-MCx | 0.495 | STN-TDM | 0.673 | GP-TDM | 0.698 | STN-MP | 0.866 | MWM-TDM | 0.688 | OC-MP | 0.887 |
| STN-GP | 0.426 | STN-OC | 0.636 | GP-OC | 0.551 | STN-MCx | 0.610 | MWM-MP | 0.429 | OC-RN | 0.855 |
| STN-SN | 0.056 | STN-SN | 0.581 | GP-CI | 0.498 | STN-TDM | 0.425 | MWM-CI | 0.401 | OC-MCx | 0.838 |
| STN-PUT | 0.052 | STN-MCx | 0.078 | GP-MCx | 0.049 | STN-SN | 0.381 | MWM-MCx | 0.284 | OC-CI | 0.804 |
| TDM-MCx | 0.720 | CI-TDM | 0.833 | STN-TDM | 0.715 | CI-MP | 0.927 | STN-TDM | 0.842 | MWM-MP | 0.977 |
| TDM-GP | 0.646 | CI-OC | 0.795 | STN-OC | 0.566 | CI-MCx | 0.672 | STN-MP | 0.543 | MWM-RN | 0.948 |
| TDM-SN | 0.108 | CI-SN | 0.738 | STN-CI | 0.511 | CI-TDM | 0.490 | STN-CI | 0.502 | MWM-MCx | 0.924 |
| TDM-PUT | 0.099 | CI-MCx | 0.131 | STN-MCx | 0.051 | CI-SN | 0.445 | STN-MCx | 0.361 | MWM-CI | 0.892 |
| MCx-GP | 0.939 | TDM-OC | 0.957 | TDM-OC | 0.829 | MP-MCx | 0.714 | TDM-MP | 0.670 | MP-RN | 0.968 |
| MCx-SN | 0.267 | TDM-SN | 0.892 | TDM-CI | 0.740 | MP-TDM | 0.512 | TDM-CI | 0.614 | MP-MCx | 0.940 |
| MCx-PUT | 0.242 | TDM-MCx | 0.155 | TDM-MCx | 0.095 | MP-SN | 0.461 | TDM-MCx | 0.450 | MP-CI | 0.906 |
| GP-SN | 0.278 | OC-SN | 0.935 | OC-CI | 0.893 | MCx-TDM | 0.817 | MP-CI | 0.908 | RN-MCx | 0.969 |
| GP-PUT | 0.252 | OC-MCx | 0.170 | OC-MCx | 0.141 | MCx-SN | 0.760 | MP-MCx | 0.714 | RN-CI | 0.935 |
| SN-PUT | 0.917 | SN-MCx | 0.194 | CI-MCx | 0.215 | TDM-SN | 0.935 | CI-MCx | 0.817 | MCx-CI | 0.968 |

**Figure S4. Anatomical lesion profile of neuropathological variables.**
The lesion profiles demonstrate differences in the patterns seen in Groups 1-4 (see manuscript) and the case with limbic predominant neuronal 4R tauopathy (LNT) pathology from Group 2 and the suspected postencephalitic parkinsonism (PEP) case in group 3. Abbreviations: GP, globus pallidus; CI, capsula interna; PUT, putamen; MCX, motor cortex; MWM, motor white matter; TDM, thalamus dorsomedial nucleus; STN, subthalamic nucleus; RN, red nucleus; MP, midbrain peduncles; SN, substantia nigra; OC, oculomotor complex.

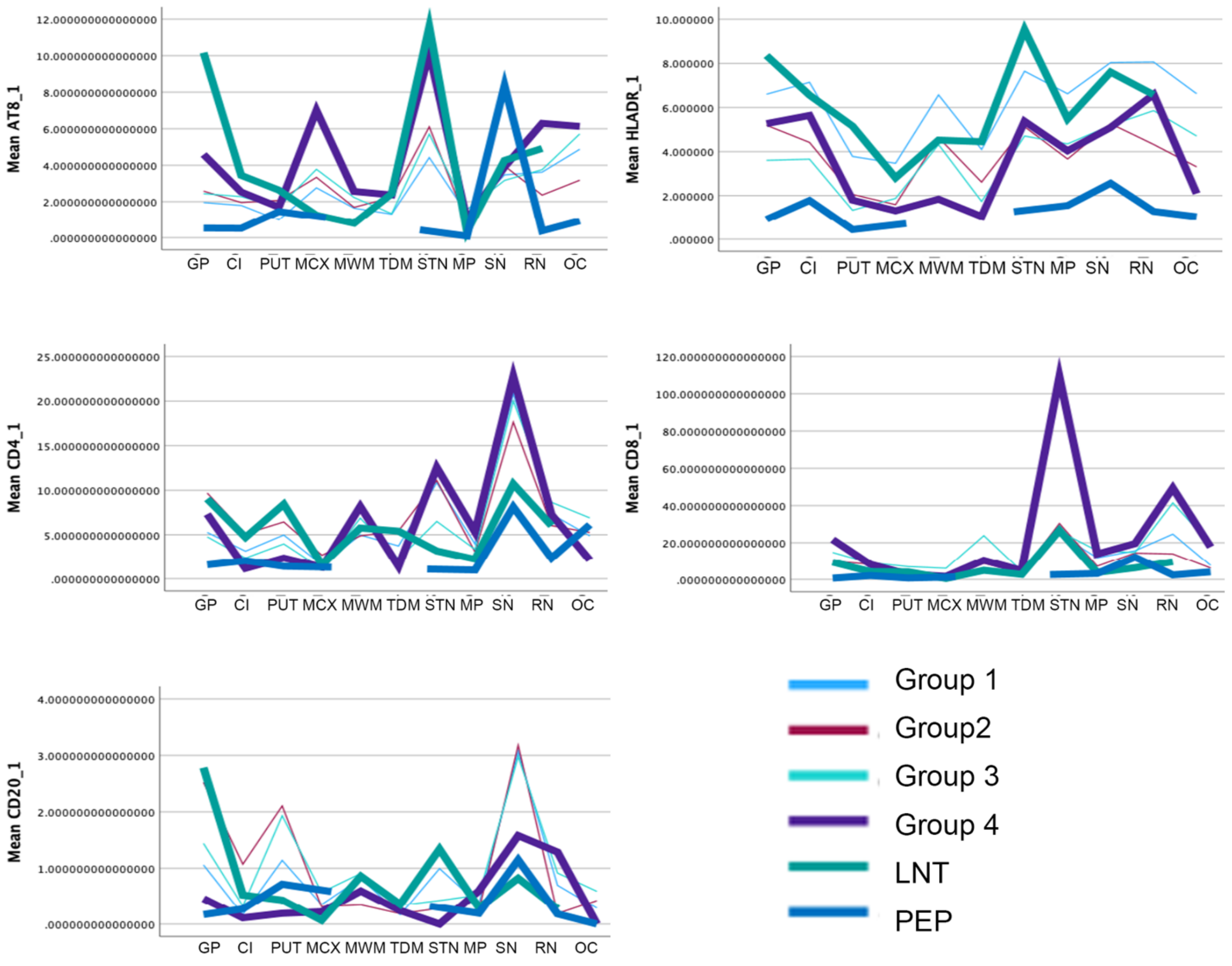

**Figure S5. Anatomical lesion profile of ratios of neuropathological variables.**
The lesion profiles demonstrate differences in the patterns seen in Groups 1-4 (see manuscript) and the case with limbic predominant neuronal 4R tauopathy (LNT) pathology from Group 2 and the suspected postencephalitic parkinsonism (PEP) case in group 3. Abbreviations: GP, globus pallidus; CI, capsula interna; PUT, putamen; MCX, motor cortex; MWM, motor white matter; TDM, thalamus dorsomedial nucleus; STN, subthalamic nucleus; RN, red nucleus; MP, midbrain peduncles; SN, substantia nigra; OC, oculomotor complex.

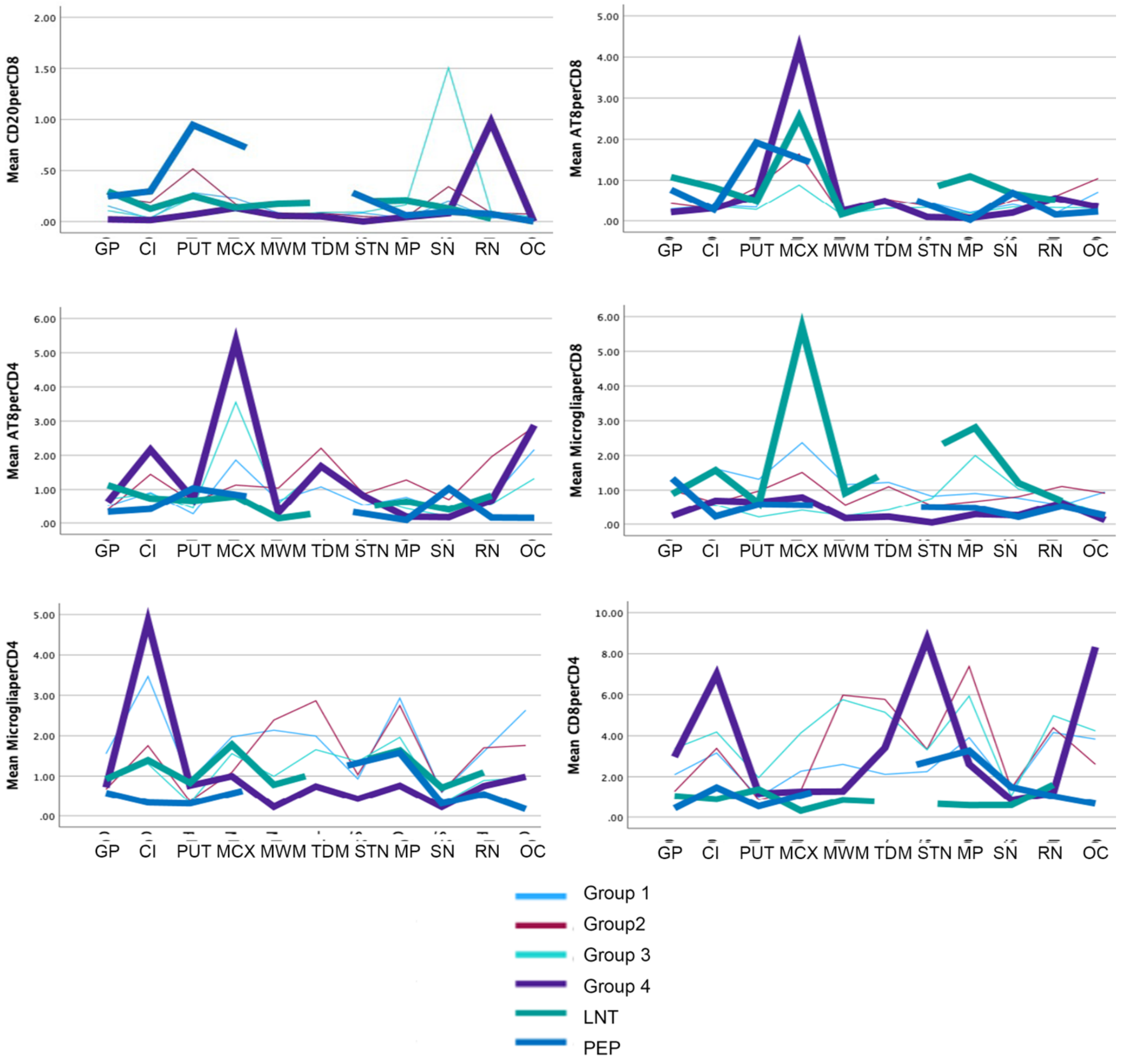
